# Increased maternal non-oxidative energy metabolism mediates association between prenatal DEHP exposure and offspring ASD symptoms: a birth cohort study

**DOI:** 10.1101/2022.06.08.22275892

**Authors:** Sarah Thomson, Katherine Drummond, Martin O’Hely, Christos Symeonides, Chitra Chandran, Toby Mansell, Richard Saffery, Peter Sly, Peter Vuillermin, Anne-Louise Ponsonby, the Barwon Infant Study Investigator Group

## Abstract

Prenatal phthalate exposure has previously been linked to the development of autism spectrum disorder (ASD). However, the underlying biological mechanisms remain unclear. We investigated whether maternal and child central carbon metabolism is involved as part of the Barwon Infant Study, a population-based birth cohort of 1074 Australian children. We estimated phthalate daily intakes using third-trimester urinary phthalate metabolite concentrations and other relevant indices. The metabolome of maternal serum in the third trimester, cord blood at birth and child plasma at 1 year were measured by nuclear magnetic resonance. We used the Small Molecule Pathway Database and principal component analysis to construct composite metabolite scores reflecting metabolic pathways. ASD symptoms at 2 and 4 years were measured by subscales of the Child Behavior Checklist and the Strengths and Difficulties Questionnaire, respectively. Multivariable linear regression analyses demonstrated (i) associations between higher prenatal di(2-ethylhexyl) phthalate (DEHP) levels and increased activity in maternal non-oxidative energy metabolism pathways, specifically non-oxidative pyruvate metabolism and the Warburg Effect, and (ii) associations between increased activity in these pathways and increased offspring ASD symptomology at 2 and 4 years of age. Mediation analyses suggested that part of the mechanism by which higher prenatal DEHP exposure influences the development of ASD symptoms in early childhood is through a maternal metabolic shift in pregnancy towards non-oxidative energy pathways, which are inefficient compared to oxidative metabolism. Interventions targeting maternal metabolic activity in pregnancy may be beneficial in reducing the potential risk to the developing fetus.

## Introduction

Autism spectrum disorder (ASD) is characterized by impaired social interaction and communication, as well as perseverative and repetitive behaviors.^1^ The etiology of ASD is not yet fully understood. However, both an adverse early environment and genetic predisposition appear important.^2^ ASD pathophysiology commences in the prenatal period^3^, an energy-demanding time of rapid brain development.^4^ ASD prevalence has risen substantially in the last two decades and is now 1–2.5%.^5^ This increase is unlikely to be explained by factors such as diagnostic changes^6^ or genetics^7^ alone. Thus, there is growing concern over the impact of environmental agents that have been epidemiologically associated with ASD,^8, 9^ and a need to understand common molecular mechanisms by which they contribute to ASD development.

Of the environmental candidates, gestational exposure to manufactured chemicals, such as phthalates, is of particular concern given increasing evidence of links to adverse early neurodevelopment.^10^ Phthalates are plasticizers used to increase the flexibility of plastics. However, as they are not bonded to the polymer, leaching occurs.^11^ They can be ingested, inhaled or dermally absorbed and major sources of exposure include food processing and packaging materials and personal care products.^12^ Almost all pregnant women in Western populations have detectable levels of phthalates in their urine.^12, 13^ Higher prenatal phthalate exposure has been associated with the development of ASD and ASD symptoms in some^14-17^ but not all^18-20^ studies. In the Barwon Infant Study, we reported that a higher combined level of four phthalates prenatally is associated with an increased likelihood of offspring ASD and ASD traits at 4 years (OR 1.55, 95% CI 1.00, 2.40; OR 1.51, 95% CI 1.20, 2.01, respectively).^21^ Given the global increase in phthalate production^22^ and the lack of success of individual-level avoidance trials thus far,^23^ there is a need to obtain a higher level of causal evidence on this issue to inform population-wide strategies. One method for achieving this is by investigating potential molecular mediators.

Both higher prenatal phthalate exposure and ASD have been associated with alterations in central carbon metabolism (referred to as ‘energy metabolism’ herein), that is, either impaired ‘oxidative metabolism’ or increased ‘non-oxidative metabolism’. Under normal conditions, oxidative processes within the mitochondria maximize energy generation **(Figure 1**, pathway 1).^24^ Under adverse conditions, like hypoxia^25, 26^ or oxidative stress^27^, activity may be diverted to less energy efficient non-oxidative processes outside the mitochondria (**Figure 1**, pathway 2).^24^ Elevated pyruvate, lactate, acetate and alanine blood levels indicate a metabolic shift towards non-oxidative processes with a more than ten-fold reduction in ATP energy production per glucose molecule compared to oxidative energy metabolism.^24^

**Figure 1.**
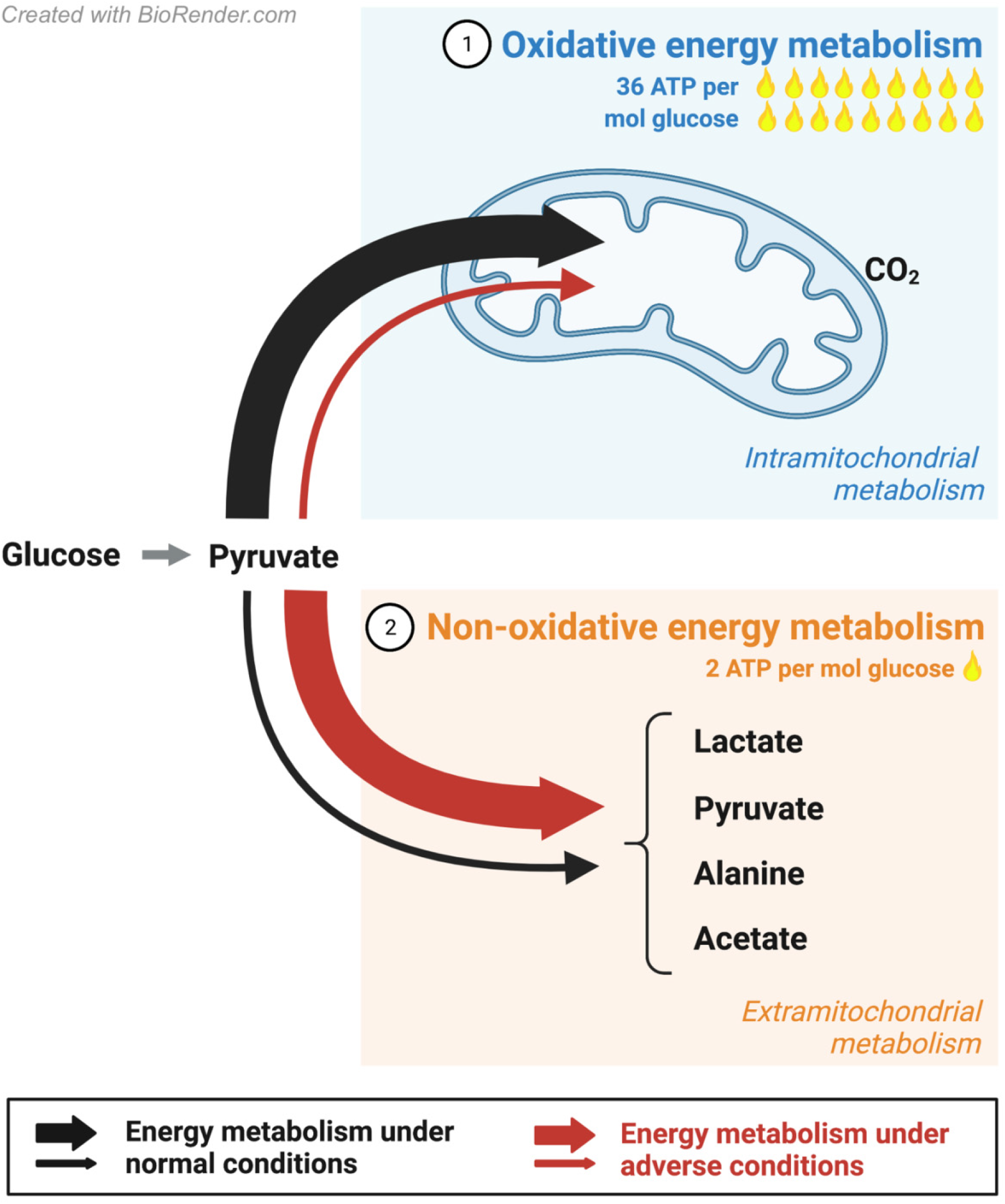
The metabolic shift towards inefficient non-oxidative energy metabolism. Normal energy metabolism typically maximizes the energy generating potential of the mitochondria through the oxidization of pyruvate (pathway 1). Metabolic shifts in central carbon metabolism can occur under adverse conditions like hypoxia or oxidative stress ^25-27^, that could be due to toxic substance exposure, or other factors. This results in the elevation of non-oxidative pathways, allowing for the diversion of carbon metabolic intermediates (pyruvate, lactate, alanine, acetate) away from the mitochondria (pathway 2) at the cost of further oxidative energy production (pathway 1). Elevated levels of these carbon metabolic intermediates in the blood or urine thus indicate a metabolic shift towards more inefficient non-oxidative energy metabolism.

Higher prenatal phthalate exposure has been positively associated with metabolic changes suggestive of reduced energy output from maternal oxidative metabolism^28, 29^ and increased non-oxidative lipid metabolism in the offspring.^30-32^ Non-oxidative carbon metabolites – pyruvate, lactate, acetate and alanine – have not, to our knowledge, been previously examined in children exposed to phthalates *in utero*.

Inefficient maternal energy metabolism in pregnancy, indicated by elevated metabolic markers of non-oxidative metabolism, has been associated with the increased occurrence of offspring ASD.^33-36^ In individuals diagnosed with ASD, energy metabolism abnormalities are common.^37, 38^ Prevalence estimates range from 30-50% for biomarkers of inefficient energy metabolism,^39^ including the elevation of serum carbon intermediates: pyruvate^40^, lactate^41^, and alanine^42^. In fact, it is estimated that 5% of individuals with ASD have classically defined mitochondrial disease^39^ compared to 0.01% of the general population.^43^

A modern causal inference technique, molecular mediation, is increasingly being employed to understand the biological mechanisms by which prenatal phthalate exposure influences adverse health outcomes.^44, 45^ However, the role of altered energy metabolism as an underlying mechanism for the link between higher prenatal phthalate levels and offspring ASD has not, to our knowledge, been evaluated. Here, we aimed to investigate (i) how phthalate exposure *in utero* associates with the mother and child’s energy metabolic profiles, (ii) how energy metabolic profiles associate with subsequent ASD symptomology in early childhood, and (iii) if the mother and/or child’s energy metabolic profiles mediate the association between prenatal phthalate exposure and ASD symptomology.

## Methods

### Cohort sample

From June 2010 to June 2013, a birth cohort of 1,074 mother–infant pairs (10 sets of twins) was recruited using an unselected antenatal sampling frame in the Barwon region of Victoria, Australia.^46^ Eligibility criteria, population characteristics and measurement details have been provided previously.^46^ The study was approved by the Barwon Health Human Research Ethics Committee (HREC 10/24) and families provided written informed consent.

### Prenatal phthalate exposure

Phthalate metabolite levels in the third trimester were measured in 842 women using a single spot urine specimen collected at 36 weeks. High-performance liquid chromatography/tandem mass spectroscopy with direct injection was performed by the Queensland Alliance for Environmental Health Science as has been outlined previously.^21^ Repeated spot specimens of monoethyl phthalate (MEP), monoisobutyl phthalate (MiBP), mono-n-butyl phthalate (MnBP), mono-(2-ethyl-5-hydroxyhexyl) phthalate (MEHHP), and mono-(2-ethyl-5-oxohexyl) phthalate (MEOHP) in the third trimester have intra-class correlation coefficients (ICC) above 0.4 in at least one of two previous studies.^47, 48^ This suggests adequate reliability of single spot specimens for capturing third-trimester phthalate exposure.

Urinary phthalate metabolite measurements were corrected for batch, specific gravity and time of day of sample collection.^21^ Phthalate estimated daily intake was then calculated accounting for maternal prenatal weight, fractional excretion of the compound, and compound-to-metabolite molecular weight ratio.^21^ The metabolites MEHHP, MEOHP and mono-(2-ethyl-5-carboxypentyl) phthalate (MECPP) were used to calculate di-(2-ethylhexyl) phthalate (DEHP) daily intake; MEP for diethyl phthalate (DEP); MiBP for diisobutyl phthalate (DiBP), and MnBP for di-n-butyl phthalate (DnBP; see **Table 1S** for abbreviations). Due to their similarity, DiBP and DnBP daily intakes were summed to make a combined daily intake measure that we will refer to as ‘DBPs’ herein. DEHP, DEP, DiBP and DnBP daily intakes were summed to make an overall composite phthalate daily intake measure. Findings expanded on our previous work^21, 49^ and considered the same phthalate measures which were log transformed to base two for analyses.

**Table 1.**
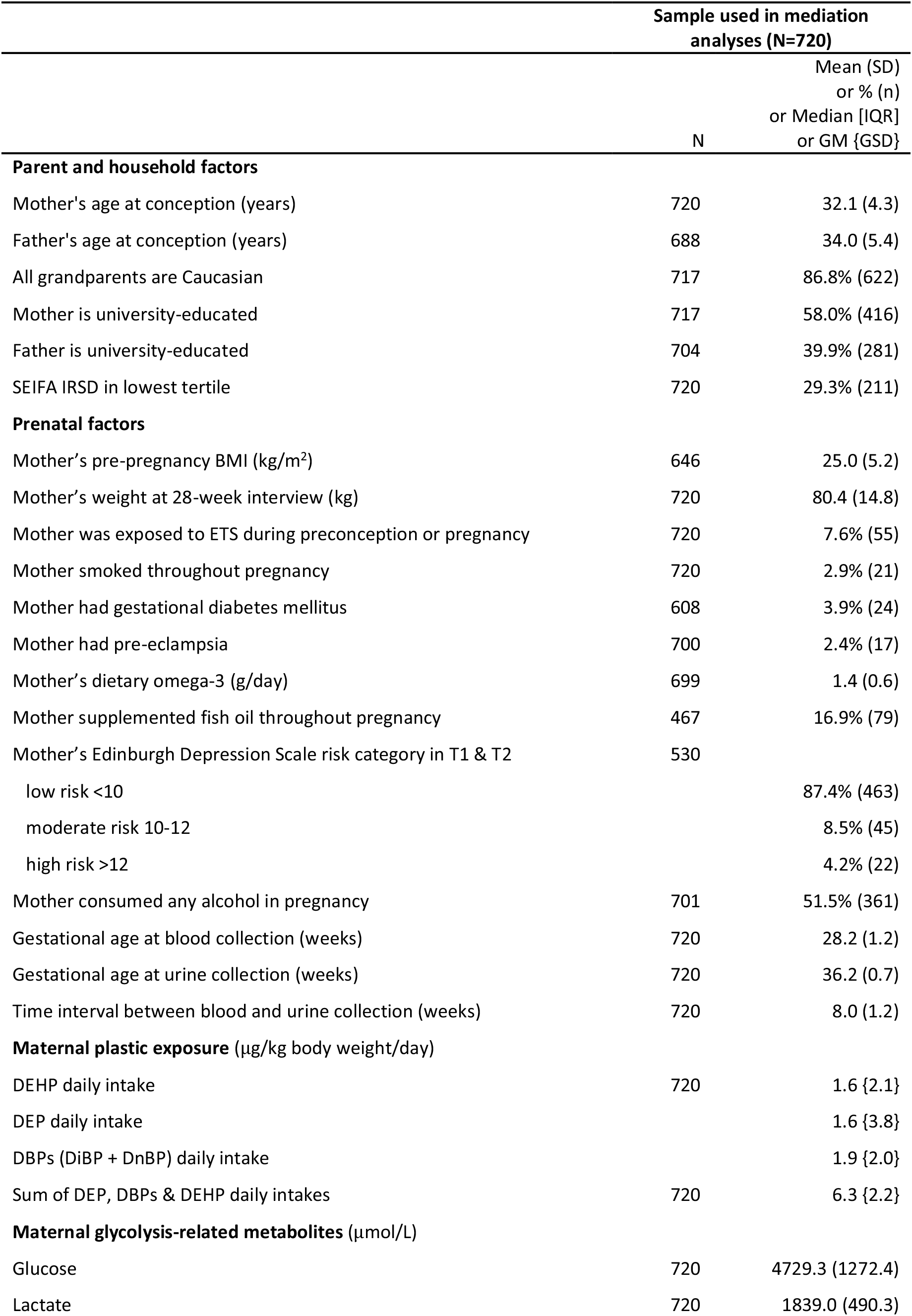
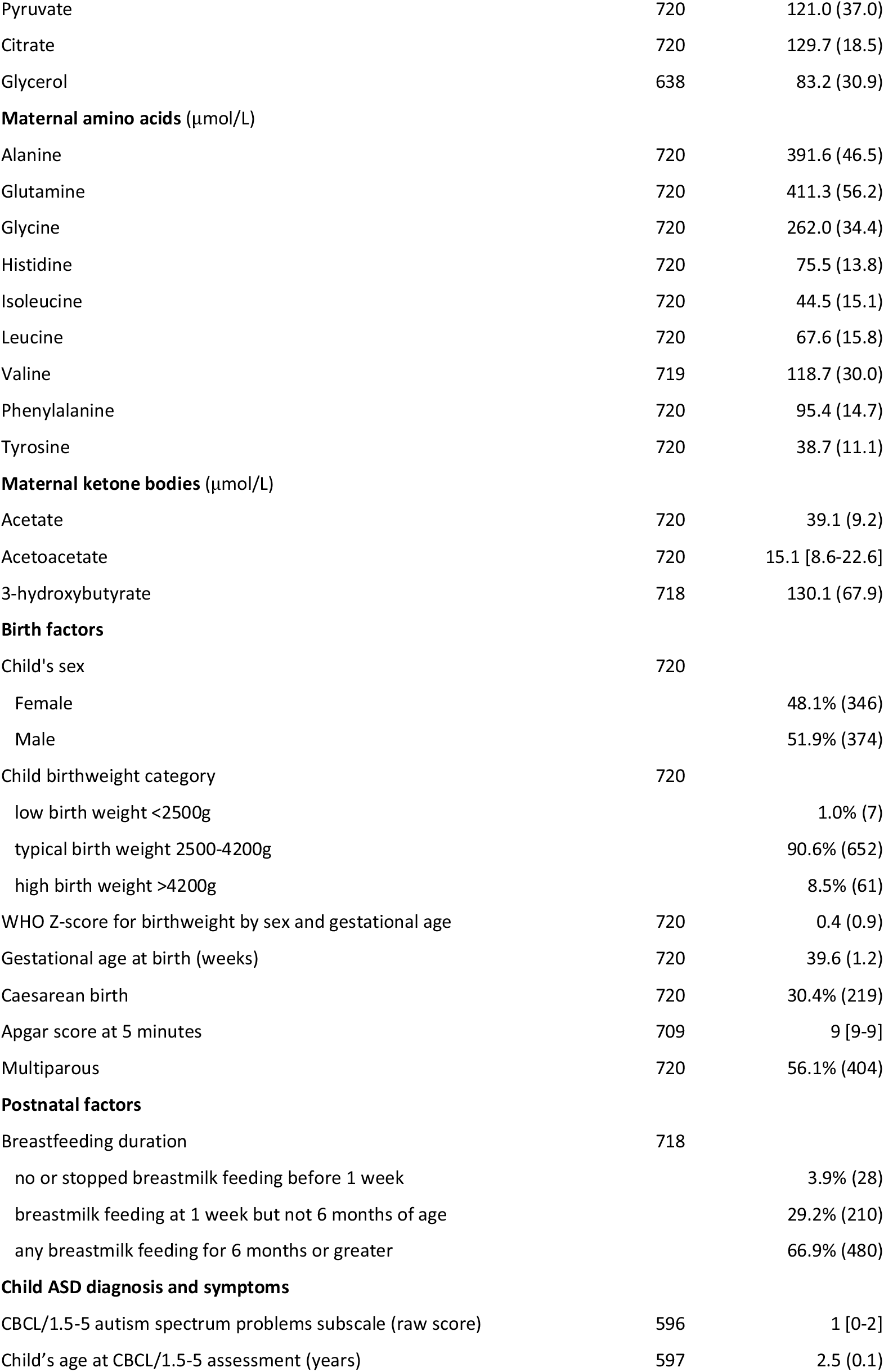
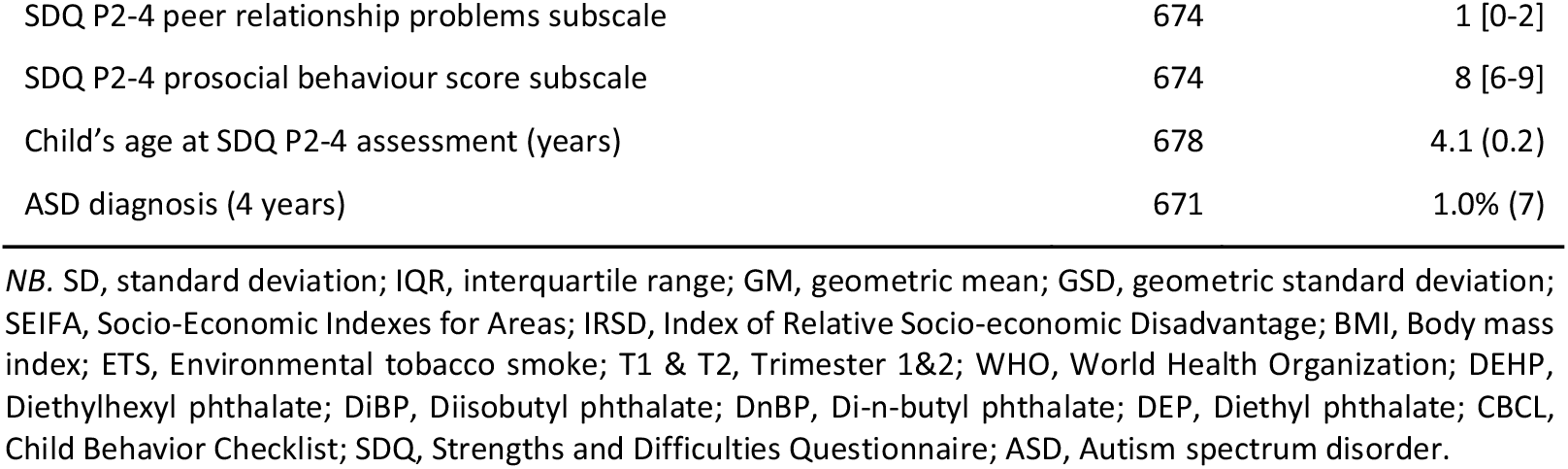
Participant characteristics.

|  | Sample used in mediation analyses (N=720) |  |
| --- | --- | --- |
|  | N | Mean (SD)<br>or % (n)<br>or Median [IQR]<br>or GM {GSD} |
| **Parent and household factors** |  |  |
| Mother's age at conception (years) | 720 | 32.1 (4.3) |
| Father's age at conception (years) | 688 | 34.0 (5.4) |
| All grandparents are Caucasian | 717 | 86.8% (622) |
| Mother is university-educated | 717 | 58.0% (416) |
| Father is university-educated | 704 | 39.9% (281) |
| SEIFA IRSD in lowest tertile | 720 | 29.3% (211) |
| **Prenatal factors** |  |  |
| Mother's pre-pregnancy BMI (kg/m<sup>2</sup>) | 646 | 25.0 (5.2) |
| Mother's weight at 28-week interview (kg) | 720 | 80.4 (14.8) |
| Mother was exposed to ETS during preconception or pregnancy | 720 | 7.6% (55) |
| Mother smoked throughout pregnancy | 720 | 2.9% (21) |
| Mother had gestational diabetes mellitus | 608 | 3.9% (24) |
| Mother had pre-eclampsia | 700 | 2.4% (17) |
| Mother's dietary omega-3 (g/day) | 699 | 1.4 (0.6) |
| Mother supplemented fish oil throughout pregnancy | 467 | 16.9% (79) |
| Mother's Edinburgh Depression Scale risk category in T1 & T2 | 530 |  |
| low risk <10 |  | 87.4% (463) |
| moderate risk 10-12 |  | 8.5% (45) |
| high risk >12 |  | 4.2% (22) |
| Mother consumed any alcohol in pregnancy | 701 | 51.5% (361) |
| Gestational age at blood collection (weeks) | 720 | 28.2 (1.2) |
| Gestational age at urine collection (weeks) | 720 | 36.2 (0.7) |
| Time interval between blood and urine collection (weeks) | 720 | 8.0 (1.2) |
| **Maternal plastic exposure (µg/kg body weight/day)** |  |  |
| DEHP daily intake | 720 | 1.6 {2.1} |
| DEP daily intake |  | 1.6 {3.8} |
| DBPs (DiBP + DnBP) daily intake |  | 1.9 {2.0} |
| Sum of DEP, DBPs & DEHP daily intakes | 720 | 6.3 {2.2} |
| **Maternal glycolysis-related metabolites (µmol/L)** |  |  |
| Glucose | 720 | 4729.3 (1272.4) |
| Lactate | 720 | 1839.0 (490.3) |
| Pyruvate | 720 | 121.0 (37.0) |
| Citrate | 720 | 129.7 (18.5) |
| Glycerol | 638 | 83.2 (30.9) |
| **Maternal amino acids (μmol/L)** |  |  |
| Alanine | 720 | 391.6 (46.5) |
| Glutamine | 720 | 411.3 (56.2) |
| Glycine | 720 | 262.0 (34.4) |
| Histidine | 720 | 75.5 (13.8) |
| Isoleucine | 720 | 44.5 (15.1) |
| Leucine | 720 | 67.6 (15.8) |
| Valine | 719 | 118.7 (30.0) |
| Phenylalanine | 720 | 95.4 (14.7) |
| Tyrosine | 720 | 38.7 (11.1) |
| **Maternal ketone bodies (μmol/L)** |  |  |
| Acetate | 720 | 39.1 (9.2) |
| Acetoacetate | 720 | 15.1 [8.6-22.6] |
| 3-hydroxybutyrate | 718 | 130.1 (67.9) |
| **Birth factors** |  |  |
| Child's sex | 720 |  |
| Female |  | 48.1% (346) |
| Male |  | 51.9% (374) |
| Child birthweight category | 720 |  |
| low birth weight <2500g |  | 1.0% (7) |
| typical birth weight 2500-4200g |  | 90.6% (652) |
| high birth weight >4200g |  | 8.5% (61) |
| WHO Z-score for birthweight by sex and gestational age | 720 | 0.4 (0.9) |
| Gestational age at birth (weeks) | 720 | 39.6 (1.2) |
| Caesarean birth | 720 | 30.4% (219) |
| Apgar score at 5 minutes | 709 | 9 [9-9] |
| Multiparous | 720 | 56.1% (404) |
| **Postnatal factors** |  |  |
| Breastfeeding duration | 718 |  |
| no or stopped breastmilk feeding before 1 week |  | 3.9% (28) |
| breastmilk feeding at 1 week but not 6 months of age |  | 29.2% (210) |
| any breastmilk feeding for 6 months or greater |  | 66.9% (480) |
| **Child ASD diagnosis and symptoms** |  |  |
| CBCL/1.5-5 autism spectrum problems subscale (raw score) | 596 | 1 [0-2] |
| Child's age at CBCL/1.5-5 assessment (years) | 597 | 2.5 (0.1) |
| SDQ P2-4 peer relationship problems subscale | 674 | 1 [0-2] |
| SDQ P2-4 prosocial behaviour score subscale | 674 | 8 [6-9] |
| Child's age at SDQ P2-4 assessment (years) | 678 | 4.1 (0.2) |
| ASD diagnosis (4 years) | 671 | 1.0% (7) |

*NB.* SD, standard deviation; IQR, interquartile range; GM, geometric mean; GSD, geometric standard deviation; SEIFA, Socio-Economic Indexes for Areas; IRSD, Index of Relative Socio-economic Disadvantage; BMI, Body mass index; ETS, Environmental tobacco smoke; T1 & T2, Trimester 1&2; WHO, World Health Organization; DEHP, Diethylhexyl phthalate; DiBP, Diisobutyl phthalate; DnBP, Di-n-butyl phthalate; DEP, Diethyl phthalate; CBCL, Child Behavior Checklist; SDQ, Strengths and Difficulties Questionnaire; ASD, Autism spectrum disorder.

### Metabolomic profiling

Metabolomic analysis was performed on non-fasting maternal serum samples collected at 28 weeks of gestation, umbilical cord serum samples, and child’s plasma samples collected at 1 year of age using the Nightingale nuclear magnetic resonance-based platform (Helsinki, Finland). Platform details can be found elsewhere.^50^ Low-molecular-weight metabolites were quantified according to Nightingale’s 2016 (maternal, child) and 2019 (cord) bioinformatics protocols.^50, 51^ Analyses were restricted to metabolites related to central carbon metabolism (amino acids, n=9; ketone bodies, n=3; glycolysis-related, n=5).

### Autism spectrum disorder: symptoms

The DSM-5-oriented autism spectrum problems subscale of the Child Behavior Checklist for Ages 1.5-5 (CBCL-ASP)^52^ and the peer relationship problems and prosocial behavior subscales of the Strengths and Difficulties Questionnaire for Ages 2-4 (SDQ-peer and SDQ-prosocial)^53^ completed by the child’s caregiver at 2-3 years and 4 years, respectively, were used as measures of ASD symptom severity. Subscale scores were calculated by summing the responses to behavioral statements (0:not true, 1:somewhat true, 2:very true). CBCL-ASP was based on 12 items (range 0-24) and SDQ-peer and SDQ-prosocial were based on 5 items (range 0-10).

In this cohort, CBCL-ASP predicted subsequent doctor-diagnosed autism with an area under the curve (AUC) of 0.92.^54^ In the literature, the CBCL/1.5-5 pervasive developmental problems scale (CBCL-PDP), SDQ-peer and SDQ-prosocial have moderate to high accuracy in distinguishing preschoolers with ASD from those who are typically developing (AUC 0.92^55^, 0.82 and 0.77^56^), respectively. CBCL-ASP has replaced CBCL-PDP to reflect changes in the DSM-5.

### Statistical methods

Pathway scores were constructed using the Small Molecule Pathway Database (SMPDB).^57^ For each central carbon metabolism pathway that contained at least three of the 17 metabolites, a principal component analysis was run on the concentrations of the metabolites in that pathway and the first principal component (PC1) was used as a composite measure. Separate multivariable linear regression models were used to estimate the associations for BIS children between (i) the phthalate measures and each of the individual and composite metabolite measures at the three timepoints (28 weeks of gestation, birth, 1 year), and (ii) the metabolite measures and each of the ASD-symptom outcomes.

Two models were run for each analysis using all available data: one with a minimal set of adjustment factors limited to child’s assigned sex at birth and process factors including gestational age at urine collection, gestational age at serum collection (prenatal timepoint only), maternal contamination of cord serum (birth timepoint only), child’s age at plasma collection (1-year timepoint only), and child’s age at behavior ratings; and another with a more extensive set of adjustment factors. For the latter, both prior knowledge and data-adaptive methods were used to (i) identify disease determinants that are independent of exposure and (ii) help separate confounders from factors that are antecedents or mediators of the putative exposure-disease associations.^58^ This data-adaptive approach is useful in low-knowledge settings where initial directed acyclic graphs may be incomplete.^58^ Focus was given to factors previously identified as being associated with ASD.^54^ Additional potential confounders were individually added to the model and the change in the estimate of the exposure-outcome association was assessed (**Table 2S**).

Metabolite pathway scores (potential mediator, *M*) that were (i) associated with any of the phthalate measures (exposure, *E*) in the regression of *M* on *E*, and (ii) with any of the ASD-symptom outcomes (outcome, *O*) in the regression of *O* on *M*, were each tested as mediators of the *E*-*O* association in formal mediation analyses. The total effect of prenatal phthalate levels on ASD symptoms was decomposed into two components: the natural direct effect (the portion of the effect that was not mediated by metabolite level) and the natural indirect effect (the portion of the effect that was mediated by metabolite level; **Figure 1S**).

Sex-specific and sex-interaction analyses were conducted. To check that the time of day of maternal serum collection did not influence the main results, sinusoidal and cosinusoidal functions of sample collection time were included as adjustment factors in the regression models. Statistical analyses were performed using Stata version 15.1 (StataCorp, USA) and R version 4.1.0 (R Foundation for Statistical Computing). Mediation analyses were implemented using the *mediation* R package.^59^

## Results

All or almost all (98%-100%) of the 842 women with phthalate measurements in the inception cohort had detectable levels of DEHP, DEP, and DBPs metabolites which varied more than 1000-fold (**Table 3S**). In the study sample (N=720 children; participant flowchart in **Figure 2;** sample characteristics in **Table 1**), the geometric mean for DEHP daily intake was 1.6μg/kg bodyweight/day (geometric SD 2.1) which is well below the current tolerable daily intake of 50μg/kg bodyweight/day in Europe.^60^ Most children exhibited few ASD symptoms and only 1% (n=7, 1 female) had an ASD diagnosis (**Table 1**). Summary statistics for metabolites are provided in **Table 1** (prenatal) and **Table 4S** (birth and 1-year).

**Figure 2.**
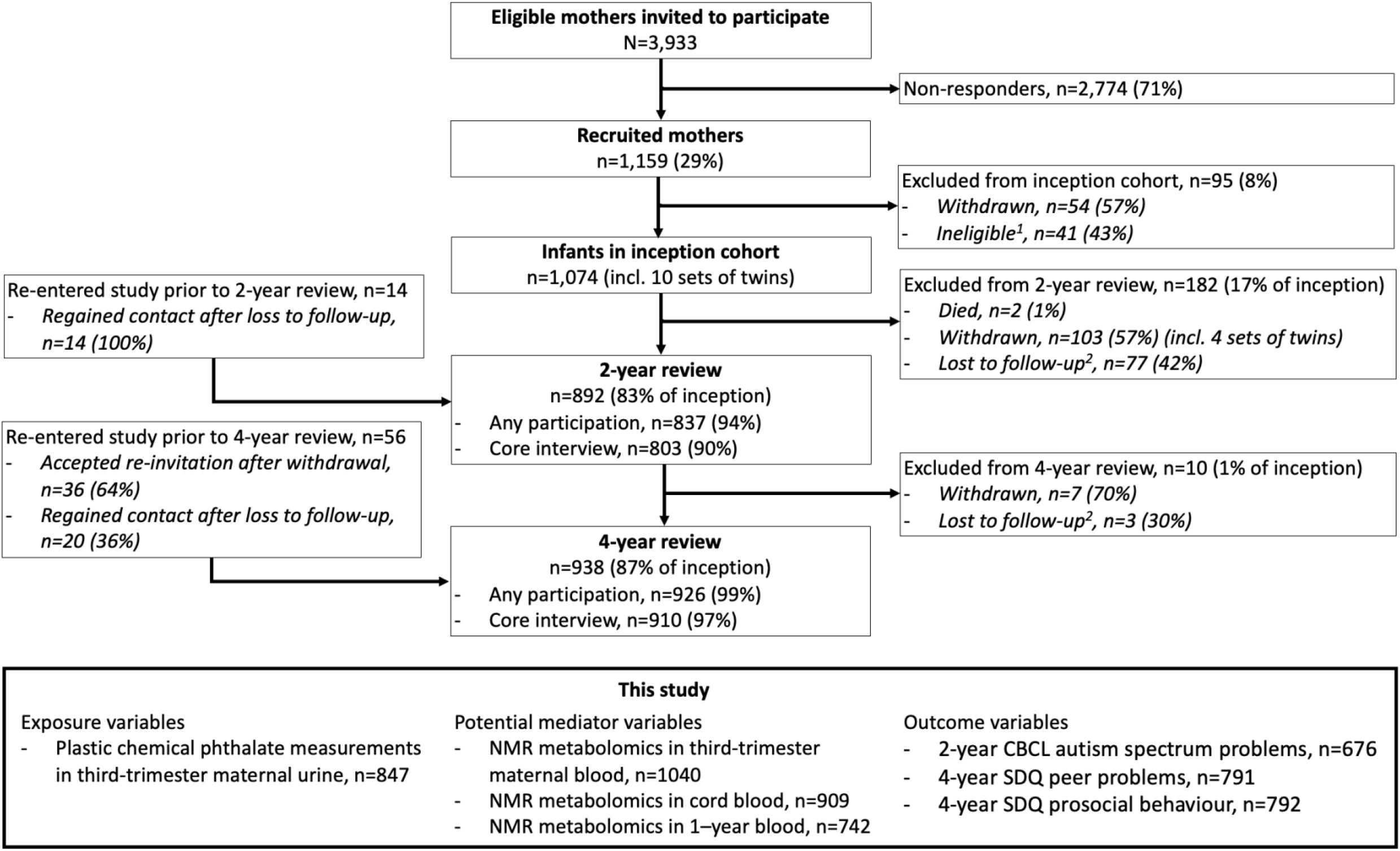
Barwon Infant Study participant flowchart. Participants included in each analysis were those with complete data on the variables of interest.

### Prenatal phthalates and maternal prenatal metabolomics

#### Higher DEHP exposure and higher pyruvate, lactate and alanine levels

Higher DEHP exposure was associated with higher levels of pyruvate and lactate (**Figure 3/2S**). The data was also somewhat compatible with a positive association between DEHP and alanine. A doubling in the daily intake of DEHP during pregnancy was associated with an estimated mean increase of 2.4μmol/L (95% CI 0.0, 4.7), 38.7μmol/L (95% CI 6.1, 71.3) and 2.8μmol/L (95% CI -0.2, 5.8) in pyruvate, lactate and alanine, respectively (**Table 5S**). When considering other phthalate measures, similar but generally weaker patterns were observed for lactate and alanine (**Figure 3S**).

**Figure 3.**
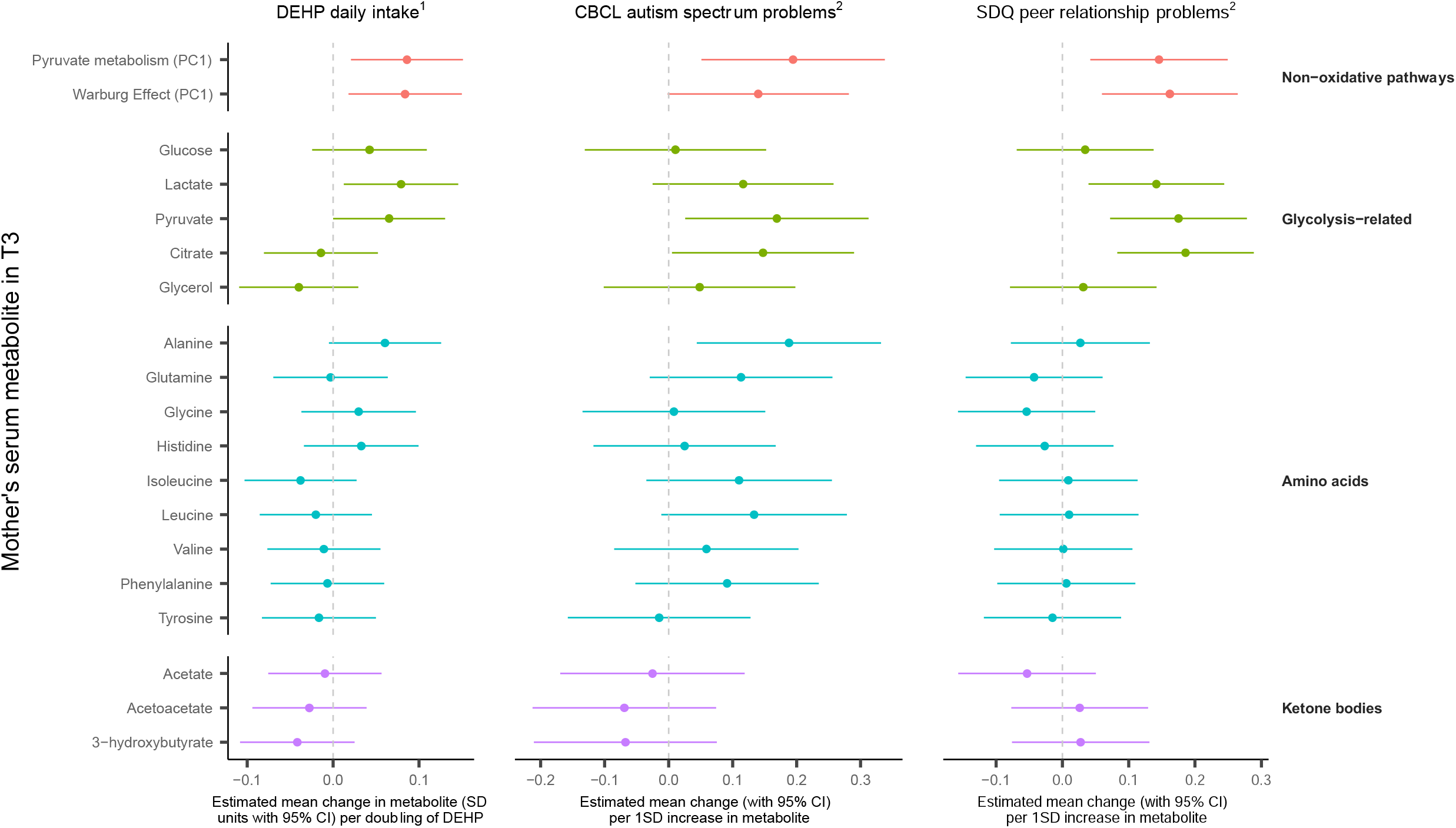
Higher prenatal maternal DEHP exposure is associated with enhanced prenatal maternal non-oxidative energy metabolism pathways. Increased activity of prenatal maternal non-oxidative energy metabolism pathways is associated with more offspring ASD symptoms. DEHP, di(2-ethylhexyl) phthalate; CBCL-ASP, Child Behavior Checklist autism spectrum problems subscale; SDQ-peer, Strengths and Difficulties Questionnaire peer relationship problems subscale; T3, Trimester 3; ^1^ Model adjusted for child’s sex, gestational age at blood collection, gestational age at urine collection, mother’s age at conception, any maternal smoking in pregnancy, maternal diet in pregnancy; ^2^ Model adjusted for child’s sex, gestational age at blood collection, child’s age at assessment of ASD symptomology, socioeconomic disadvantage, maternal multiparity.

#### Higher DEHP exposure associated with higher non-oxidative energy pathway scores

In non-oxidative pyruvate metabolism, pyruvate is converted to lactate, acetate and alanine and our Non-Oxidative Pyruvate Metabolism Score (NOPMS) captured 45% of the overall variability. In the Warburg Effect, glucose is converted to pyruvate which is then converted to lactate and our Warburg Effect Metabolism Score (WEMS) captured 57% of the overall variability. A doubling in the daily intake of DEHP during pregnancy was associated with an estimated mean increase of 0.09 SD units (95% CI 0.02, 0.15) and 0.08 SD units (95% CI 0.02, 0.15) in the NOPMS and WEMS, respectively (**Figure 3/2S**). Similar but weaker patterns were observed for the composite phthalate measure (**Figure 3S**). However, for DEP and the DBPs, the data had low compatibility with any association being present which suggests the findings for the composite phthalate measure were primarily driven by DEHP (**Figure 3S**).

### Maternal prenatal metabolomics and offspring ASD symptoms

#### Higher pyruvate, lactate, citrate and alanine levels associated with higher ASD symptom scores

For a 1 SD elevation in pyruvate, citrate and alanine, the estimated mean increase in CBCL-ASP at 2 years was 0.17 points (95% CI 0.03, 0.31), 0.15 points (95% CI 0.01, 0.29), and 0.19 points (95% CI 0.04, 0.33), respectively (**Figure 3/2S**). For a 1 SD elevation in pyruvate, citrate and lactate, the estimated mean increase in SDQ-peer problems at 4 years was 0.18 points (95% CI 0.07, 0.28), 0.19 points (95% CI 0.08, 0.29), and 0.14 points (95% CI 0.04, 0.24), respectively (**Figure 3/2S**). See **Table 5S** for unscaled regression estimates.

#### Higher non-oxidative energy pathway scores associated with higher ASD symptom scores

The estimated mean increase in CBCL-ASP score at 2 years for a 1 SD elevation in NOPMS and WEMS was 0.19 points (95% CI 0.05, 0.34) and 0.14 points (95% CI 0.00, 0.28), respectively (**Figure 3/2S**). Per 1 SD elevation in NOPMS and WEMS, the estimated mean increase in SDQ-peer problems score at 4 years was 0.15 points (95% CI 0.04, 0.25) and 0.16 points (95% CI 0.06, 0.26), respectively (**Figure 3/2S**).

### Metabolomics in cord blood and child’s blood at 1 year

There were no consistent findings across or within the *exposure-mediator* and *mediator*-*outcome* models at the perinatal (cord serum) or postnatal (1 year child’s blood) time points (**Figure 4S-7S**).

**Figure 4.**
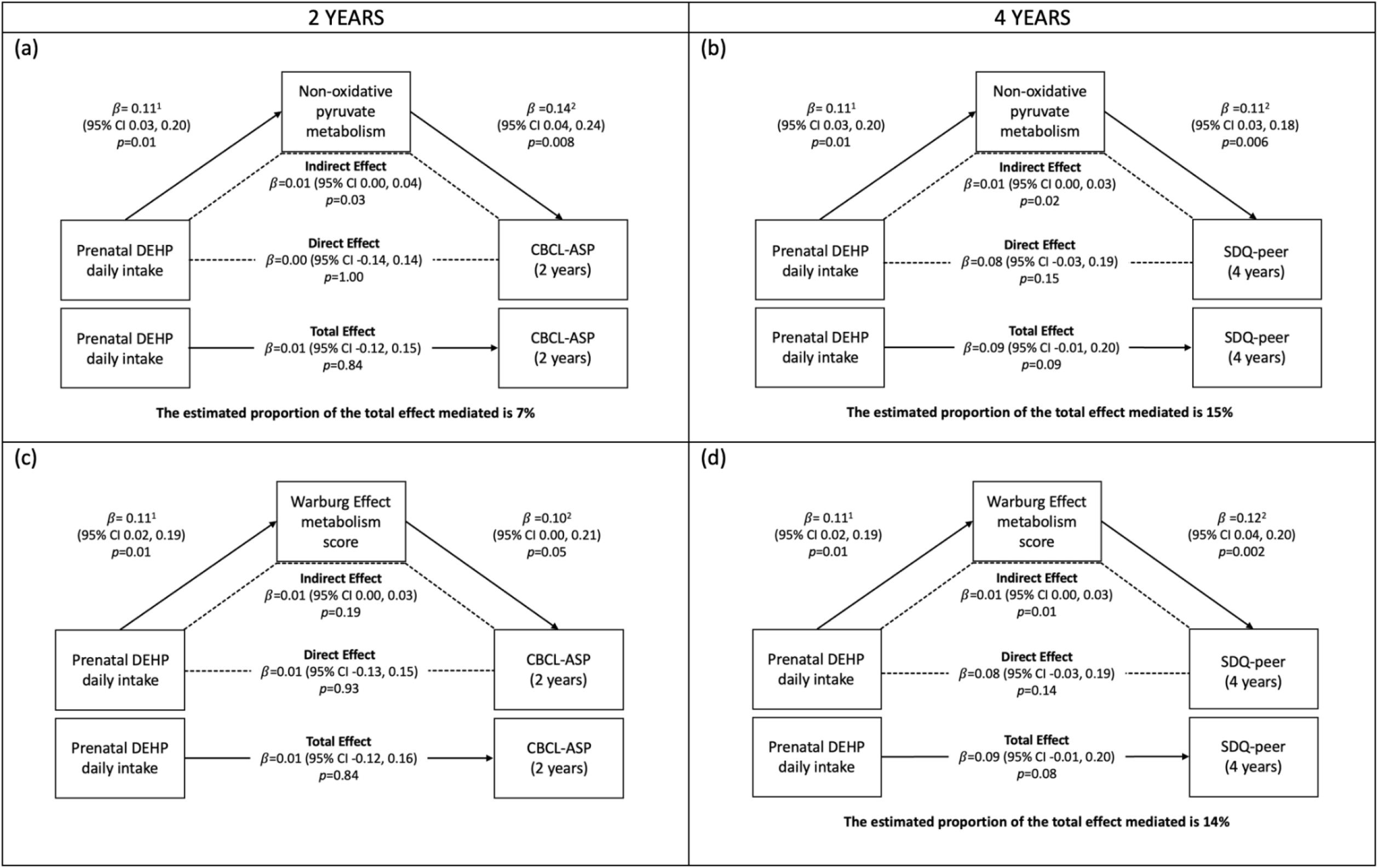
Prenatal maternal non-oxidative pyruvate metabolism (a and b) and Warburg Effect metabolism (d) are partial mediators of the association between prenatal maternal DEHP exposure and offspring ASD symptomology in early childhood. Models with metabolism pathway score as the dependent variable are adjusted for child’s sex, gestational age at blood collection, gestational age at urine collection, mother’s age at conception, any maternal smoking in pregnancy, and maternal diet during pregnancy; models with an ASD symptoms measure as the dependent variable are adjusted for child’s sex, gestational age at blood collection, child’s age at ASD symptom questionnaire, socioeconomic disadvantage, and maternal multiparity; DEHP, di(2-ethylhexyl) phthalate; CBCL-ASP, Child Behavior Checklist autism spectrum problems subscale; SDQ-peer, Strengths and Difficulties Questionnaire peer relationship problems subscale; ^1^ Estimated increase in metabolite (mmol/L) per doubling of prenatal DEHP daily intake; ^2^ Estimated increase in ASD symptom score per mmol/L increase in metabolite measure.

### Mediation analyses

#### Increased maternal non-oxidative energy metabolism in pregnancy partly mediates positive association between prenatal DEHP exposure and ASD symptoms

The indirect effect estimates suggest that elevations in maternal NOPMS and WEMS each partially mediate the association between higher prenatal DEHP exposure and increased ASD symptomology in early childhood (**Figure 4a, 4b, 4d**). That is, the data are compatible with the hypothesis that higher levels of prenatal DEHP exposure increase maternal non-oxidative energy metabolism during pregnancy, which in turn increases the likelihood of offspring ASD symptoms. However, there was a lower degree of compatibility when WEMS and CBCL-ASP were considered as mediator and outcome, respectively (**Figure 4c**).

### Additional analyses

Generally, differences by sex were not detected (**Figure 8S**). There was weak evidence of an interaction between sex and the composite phthalate daily intake measure, suggesting the association between phthalate exposure and increased non-oxidative energy metabolism is stronger in mothers carrying male compared to female fetuses. Larger sample sizes are needed in future work to further investigate sex differences. Including sinusoidal and cosinusoidal functions of time of maternal serum collection as adjustment factors in the regression models did not materially change the findings.

## Discussion

Using causal mediation analyses, this study is the first to report that a metabolic shift in maternal energy metabolism in pregnancy partially underlies the association between higher prenatal DEHP exposure and increased offspring ASD symptomology in early childhood. This shift is towards extra-mitochondrial non-oxidative pathways – non-oxidative pyruvate metabolism and the Warburg Effect – that are far less efficient at generating energy than intramitochondrial oxidative phosphorylation.

In this study, higher prenatal DEHP exposure was associated with elevated lactate, pyruvate, NOPMS and WEMS in maternal serum during pregnancy. Similar positive associations between phthalate levels and non-oxidative metabolites (lactate and pyruvate) have been reported in maternal prenatal urine samples.^28, 29^ DEHP exposure also causes an accumulation of lactate, suggestive of increased non-oxidative metabolism, in various cell types *in vitro* (cardiomyocytes^61^, adipocytes^62, 63^ and Sertoli cells^64^) and muscle tissue *in vivo*^65^, and is linked to disrupted regulation of enzymes along central carbon metabolism pathways (**Figure 9S**).

DEHP could act directly or indirectly to cause a metabolic shift towards non-oxidative energy metabolism in pregnant women. DEHP and its metabolites may enhance the activity of non-oxidative energy metabolism pathways *directly* by binding to or altering the activity of regulators of non-oxidative metabolism (for example, the mitochondrial pyruvate carrier complex or peroxisome proliferator-activated receptors^66, 67^). DEHP could also cause this metabolic shift *indirectly*. For instance, DEHP exposure is associated with an elevation in cellular oxidative stress,^68^ which is linked to a subsequent increase in carbon intermediates.^27, 69^ DEHP may also contribute to mitochondrial dysfunction,^70, 71^ which diminishes cellular energy supply and increases the utilization of non-oxidative energy metabolism pathways. Alternatively, a DEHP-induced elevation of non-oxidative metabolites could be due to a high glycolytic rate that strains the oxidative capacity of the mitochondria^72^; however, there is inconsistent evidence of hyperactive glycolysis after DEHP exposure.^71^ Overall, DEHP, through several possible direct or indirect actions, could increase the utilization of non-oxidative energy pathways.

A metabolic shift towards maternal non-oxidative energy metabolism during pregnancy could arise through multiple underlying biological processes with known links to ASD. Oxidative stress, reported to be induced by DEHP exposure^44^, is more likely to be chronic during pregnancy in mothers whose children develop ASD^33^ and greater oxidative stress is associated with the generation of non-oxidative energy metabolites.^72, 73^ Similarly, underlying mitochondrial dysfunction increases metabolic diversion to non-oxidative pathways^73, 74^ and mothers with disrupted mitochondrial function during pregnancy are also more likely to have children who develop ASD.^39^ The existing links in the literature between these two biological mechanisms^39^ that both increase non-oxidative energy metabolism and offspring ASD risk further support our finding that DEHP-induced disrupted energy metabolism contributes to the development of offspring ASD symptomology.

This is the first report that elevated non-oxidative energy metabolism during pregnancy is associated with increased offspring ASD symptomology at both 2 and 4 years. A shift toward non-oxidative metabolism provides a possible unifying mechanism for a variety of prenatal environmental exposures reported to increase the risk of offspring ASD. For instance, studies of pregnant women with conditions known to increase the risk of offspring ASD diagnosis (e.g. gestational diabetes^75^, obesity^76^, psychological stress^77^ and preeclampsia^78^) incidentally report an elevation of non-oxidative energy metabolites during pregnancy.^79^ Furthermore, women with deviations in central carbon metabolism during pregnancy after exposure to high levels of air pollution are more likely to have offspring with ASD.^36^ Similarly, mothers exposed to chemicals known to increase non-oxidative metabolites (e.g. valproate^80^ and dexamethasone^81^) are also more likely to have a child diagnosed with ASD.^82^ Rodent studies have demonstrated that manipulations of maternal prenatal energy metabolism can result in offspring structural brain abnormalities that are a hallmark of ASD.^83-85^

Strengths of the study include comprehensive data on prenatal environmental measures, unique serial carbon metabolomic indices and the use of previously validated ASD symptom scales from ages two to four years in a large study sample, providing the first setting, to our knowledge, where these factors have been examined together. The use of pathway scores allowed us to evaluate the composite impact of related metabolites in a biologically defined pathway. The highly dimensioned cohort enabled an evaluation of non-causal and causal factors, and the findings of regression analyses with the minimal adjustment set persisted after inclusion of confounders. The use of modern causal inference methods allowed us to show molecular mediation. Other causal features such as plausibility and dose response were also present.

It is likely that that elevated non-oxidative energy metabolism is a marker of a closely related biological primary cause such as impaired mitochondrial oxidative phosphorylation. However, acetyl CoA and other relevant measures were not available to provide a readout on oxidative energy metabolism. Further, this project did not perform functional assays such as assessing maternal mitochondrial function in pregnancy (e.g. using Seahorse XFe96 Analyzer^86^) and further work on this should be conducted. The sample was not large enough to study ASD diagnosis as a main outcome or interrogate differences by sex. Given an ASD prevalence here of 1%, a cohort sample of 6,000 would have been required to provide 60 ASD cases and comparator children with highly dimensioned data including serial metabolomics. Such cohorts do not, to our knowledge, currently exist. Only single urine and blood samples were collected from the mother during pregnancy. However, past work has found repeat phthalate measures in pregnancy to be moderately reproducible,^47, 48^ and we have further accounted for some of the features (time of day, maternal weight) associated with variability. Maternal blood lactate and pyruvate levels are also relatively consistent across trimester 3.^87, 88^ Further, error in measurement introduced by single samples would tend to bias estimates towards the null. While, for most mothers, phthalate urinary metabolites were measured later in trimester 3 than the maternal serum metabolites, adjustment for this time interval had little effect and the strength of association between phthalate levels and pathway scores did not materially differ by time interval between serum and urine measures. Finally, serum and plasma were non-fasting samples. An assessment of the impact of time of maternal blood collection showed it had little effect on the results. Our results highlight the important role of the prenatal environment in ASD causation and suggest that part of the mechanism by which prenatal exposure to DEHP influences offspring ASD symptom development is through factors related to a metabolic shift in maternal energy metabolism in pregnancy. A shift towards non-oxidative metabolism, which is inefficient compared to oxidative metabolism, may be a common biological response to a variety of prenatal environmental exposures that also increase the risk of offspring ASD. Thus, strategies of managing or preventing deviation toward non-oxidative energy metabolism in the mother during pregnancy may be beneficial in reducing the potential risk to the developing fetus.

## Supporting information

COI Disclosure

STROBE Checklist

## Data Availability

Access to Barwon Infant Study (BIS) data including all data used in this paper can be requested through the BIS Steering Committee by contacting the corresponding author. Requests to access cohort data are considered on scientific and ethical grounds and, if approved, provided under collaborative research agreements. De-identified cohort data can be provided in Stata or CSV format. Additional project information, including cohort data description and access procedure, is available at the project website https://www.barwoninfantstudy.org.au.

## Acknowledgments

The authors thank the BIS participants for the generous contribution they have made to this project. The authors also thank current and past staff for their efforts in recruiting and maintaining the cohort and in obtaining and processing the data and biospecimens. The establishment work and infrastructure for the BIS was provided by the Murdoch Children’s Research Institute, Deakin University and Barwon Health. Subsequent funding was secured from the Minderoo Foundation, the European Union’s Horizon 2020 research and innovation programme (ENDpoiNTs: No 825759), National Health and Medical Research Council of Australia (NHMRC), The Shepherd Foundation, The Jack Brockhoff Foundation, the Scobie & Claire McKinnon Trust, the Shane O’Brien Memorial Asthma Foundation, the Our Women Our Children’s Fund Raising Committee Barwon Health, the Rotary Club of Geelong, the Ilhan Food Allergy Foundation, GMHBA, Vanguard Investments Australia Ltd, and the Percy Baxter Charitable Trust, Perpetual Trustees. In-kind support was provided by the Cotton On Foundation and CreativeForce. The study sponsors were not involved in the collection, analysis, and interpretation of data; writing of the report; or the decision to submit the report for publication. Research at Murdoch Children’s Research Institute is supported by the Victorian Government’s Operational Infrastructure Support Program.

The other members of the BIS Investigator Group are Mimi LK Tang, Amy Loughman, Lawrence Gray, Sarath Ranganathan, and David Burgner. We thank Terry Dwyer and Katie Allen for their past work as foundation investigators and John Carlin for statistical advice in the Barwon Infant Study.

## References

1. Jonsson U, Alaie I, Löfgren Wilteus A, Zander E, Marschik PB, Coghill D et al. Annual Research Review: Quality of life and childhood mental and behavioural disorders–a critical review of the research. Journal of child psychology and psychiatry 2017; 58(4): 439–469.

2. Hallmayer J, Cleveland S, Torres A, Phillips J, Cohen B, Torigoe T et al. Genetic Heritability and Shared Environmental Factors Among Twin Pairs With Autism. Archives of General Psychiatry 2011; 68(11): 1095–1102.

3. Connors SL, Levitt P, Matthews SG, Slotkin TA, Johnston MV, Kinney HC et al. Fetal mechanisms in neurodevelopmental disorders. Pediatr Neurol 2008; 38(3): 163–176.

4. Steiner P. Brain Fuel Utilization in the Developing Brain. Annals of Nutrition and Metabolism 2020; 75(Suppl 1): 8.

5. Atladottir HO, Gyllenberg D, Langridge A, Sandin S, Hansen SN, Leonard H et al. The increasing prevalence of reported diagnoses of childhood psychiatric disorders: a descriptive multinational comparison. European child & adolescent psychiatry 2015; 24(2): 173–183.

6. Hansen SN, Schendel DE, Parner ET. Explaining the increase in the prevalence of autism spectrum disorders: the proportion attributable to changes in reporting practices. JAMA pediatrics 2015; 169(1): 56–62.

7. Herbert MR, Russo JP, Yang S, Roohi J, Blaxill M, Kahler SG et al. Autism and environmental genomics. Neurotoxicology 2006; 27(5): 671–684.

8. Bölte S, Girdler S, Marschik PB. The contribution of environmental exposure to the etiology of autism spectrum disorder. Cell Mol Life Sci 2019; 76(7): 1275–1297.

9. Rossignol DA, Genuis SJ, Frye RE. Environmental toxicants and autism spectrum disorders: a systematic review. Transl Psychiatry 2014; 4(2): e360.

10. Bennett D, Bellinger DC, Birnbaum LS, Bradman A, Chen A, Cory-Slechta DA et al. Project TENDR: Targeting Environmental Neuro-Developmental Risks The TENDR Consensus Statement. Environ Health Perspect 2016; 124(7): A118–122.

11. Hahladakis JN, Velis CA, Weber R, Iacovidou E, Purnell P. An overview of chemical additives present in plastics: migration, release, fate and environmental impact during their use, disposal and recycling. vol. 3442018, pp 179–199.

12. Sugeng EJ, Symeonides C, O’Hely M, Vuillermin P, Sly PD, Vijayasarathy S et al. Predictors with regard to ingestion, inhalation and dermal absorption of estimated phthalate daily intakes in pregnant women: The Barwon infant study. Environ Int 2020; 139: 105700.

13. Shin HM, Dhar U, Calafat AM, Nguyen V, Schmidt RJ, Hertz-Picciotto I. Temporal Trends of Exposure to Phthalates and Phthalate Alternatives in California Pregnant Women during 2007-2013: Comparison with Other Populations. Environ Sci Technol 2020; 54(20): 13157– 13166.

14. Larsson M, Weiss B, Janson S, Sundell J, Bornehag C-G. Associations between indoor environmental factors and parental-reported autistic spectrum disorders in children 6–8 years of age. Neurotoxicology 2009; 30(5): 822–831.

15. Miodovnik A, Engel SM, Zhu C, Ye X, Soorya LV, Silva MJ et al. Endocrine disruptors and childhood social impairment. Neurotoxicology 2011; 32(2): 261–267.

16. Oulhote Y, Lanphear B, Braun JM, Webster GM, Arbuckle TE, Etzel T et al. Gestational exposures to phthalates and folic acid, and autistic traits in canadian children. Environmental health perspectives 2020; 128(2): 027004.

17. Haggerty DK, Strakovsky RS, Talge NM, Carignan CC, Glazier-Essalmi AN, Ingersoll BR et al. Prenatal phthalate exposures and autism spectrum disorder symptoms in low-risk children. Neurotoxicology and Teratology 2021; 83: 106947.

18. Braun JM, Kalkbrenner AE, Just AC, Yolton K, Calafat AM, Sjödin A et al. Gestational exposure to endocrine-disrupting chemicals and reciprocal social, repetitive, and stereotypic behaviors in 4-and 5-year-old children: the HOME study. Environmental health perspectives 2014; 122(5): 513–520.

19. Shin H-M, Schmidt RJ, Tancredi D, Barkoski J, Ozonoff S, Bennett DH et al. Prenatal exposure to phthalates and autism spectrum disorder in the MARBLES study. Environmental Health 2018; 17(1): 1–14.

20. Radke EG, Braun JM, Nachman RM, Cooper GS. Phthalate exposure and neurodevelopment: A systematic review and meta-analysis of human epidemiological evidence. Environment International 2020; 137: 105408.

21. Ponsonby A-L, Symeonides C, Saffery R, Mueller JF, O’Hely M, Sly PD et al. Prenatal phthalate exposure, oxidative stress-related genetic vulnerability and early life neurodevelopment: a birth cohort study. Neurotoxicology 2020; 80: 20–28.

22. Prasad B. Phthalate pollution: environmental fate and cumulative human exposure index using the multivariate analysis approach. Environmental Science: Processes & Impacts 2021.

23. Sathyanarayana S, Alcedo G, Saelens BE, Zhou C, Dills RL, Yu J et al. Unexpected results in a randomized dietary trial to reduce phthalate and bisphenol A exposures. J Expo Sci Environ Epidemiol 2013; 23(4): 378–384.

24. Lubert S, John LT, Jeremy MB. Biochemistry fifth edition. 2015.

25. Koziel A, Jarmuszkiewicz W. Hypoxia and aerobic metabolism adaptations of human endothelial cells. Pflugers Arch 2017; 469(5-6): 815–827.

26. Chicco AJ, L. CH, Gnaiger E, Dreyer HC, Muyskens JB, D’Alessandro A et al. Adaptive remodeling of skeletal muscle energy metabolism in high-altitude hypoxia: Lessons from AltitudeOmics. J Biol Chem 2018; 293(18): 6659–6671.

27. Liu X, Cooper DE, Cluntun AA, Warmoes MO, Zhao S, Reid MA et al. Acetate Production from Glucose and Coupling to Mitochondrial Metabolism in Mammals. Cell 2018; 175(2): 502–513.e513.

28. Maitre L, Robinson O, Martinez D, Toledano MB, Ibarluzea J, Marina LS et al. Urine metabolic signatures of multiple environmental pollutants in pregnant women: an exposome approach. Environmental science & technology 2018; 52(22): 13469–13480.

29. Zhou M, Ford B, Lee D, Tindula G, Huen K, Tran V et al. Metabolomic markers of phthalate exposure in plasma and urine of pregnant women. Frontiers in public health 2018; 6: 298.

30. Kupsco A, Wu H, Calafat AM, Kioumourtzoglou MA, Tamayo-Ortiz M, Pantic I et al. Prenatal maternal phthalate exposures and child lipid and adipokine levels at age six: A study from the PROGRESS cohort of Mexico City. Environ Res 2021; 192: 110341.

31. Perng W, Watkins DJ, Cantoral A, Mercado-García A, Meeker JD, Téllez-Rojo MM et al. Exposure to phthalates is associated with lipid profile in peripubertal Mexican youth. Environ Res 2017; 154: 311–317.

32. Vafeiadi M, Myridakis A, Roumeliotaki T, Margetaki K, Chalkiadaki G, Dermitzaki E et al. Association of Early Life Exposure to Phthalates With Obesity and Cardiometabolic Traits in Childhood: Sex Specific Associations. Front Public Health 2018; 6: 327.

33. Hollowood K, Melnyk S, Pavliv O, Evans T, Sides A, Schmidt RJ et al. Maternal metabolic profile predicts high or low risk of an autism pregnancy outcome. Research in autism spectrum disorders 2018; 56: 72–82.

34. Lyall K, Schmidt RJ, Hertz-Picciotto I. Maternal lifestyle and environmental risk factors for autism spectrum disorders. Int J Epidemiol 2014; 43(2): 443–464.

35. Egorova O, Myte R, Schneede J, Hägglöf B, Bölte S, Domellöf E et al. Maternal blood folate status during early pregnancy and occurrence of autism spectrum disorder in offspring: a study of 62 serum biomarkers. Mol Autism 2020; 11(1): 7.

36. Kim JH, Yan Q, Uppal K, Cui X, Ling C, Walker DI et al. Metabolomics analysis of maternal serum exposed to high air pollution during pregnancy and risk of autism spectrum disorder in offspring. Environ Res 2021; 196: 110823.

37. Cheng N, Rho JM, Masino SA. Metabolic Dysfunction Underlying Autism Spectrum Disorder and Potential Treatment Approaches. Frontiers in molecular neuroscience 2017; 10: 34–34.

38. Dhillon S, Hellings JA, Butler MG. Genetics and mitochondrial abnormalities in autism spectrum disorders: a review. Current genomics 2011; 12(5): 322–332.

39. Frye RE, Cakir J, Rose S, Palmer RF, Austin C, Curtin P et al. Mitochondria May Mediate Prenatal Environmental Influences in Autism Spectrum Disorder. Journal of Personalized Medicine 2021; 11(3): 218.

40. Hassan MH, Desoky T, Sakhr HM, Gabra RH, Bakri AH. Possible metabolic alterations among autistic male children: clinical and biochemical approaches. Journal of Molecular Neuroscience 2019; 67(2): 204–216.

41. Correia C, Coutinho AM, Diogo L, Grazina M, Marques C, Miguel T et al. Brief report: High frequency of biochemical markers for mitochondrial dysfunction in autism: no association with the mitochondrial aspartate/glutamate carrier SLC25A12 gene. Journal of autism and Developmental Disorders 2006; 36(8): 1137–1140.

42. Orozco JS, Hertz-Picciotto I, Abbeduto L, Slupsky CM. Metabolomics analysis of children with autism, idiopathic-developmental delays, and Down syndrome. Transl Psychiatry 2019; 9(1): 243.

43. Skladal D, Halliday J, Thorburn DR. Minimum birth prevalence of mitochondrial respiratory chain disorders in children. Brain 2003; 126(8): 1905–1912.

44. Ferguson KK, Chen YH, VanderWeele TJ, McElrath TF, Meeker JD, Mukherjee B. Mediation of the Relationship between Maternal Phthalate Exposure and Preterm Birth by Oxidative Stress with Repeated Measurements across Pregnancy. Environ Health Perspect 2017; 125(3): 488–494.

45. England-Mason G, Grohs MN, Reynolds JE, MacDonald A, Kinniburgh D, Liu J et al. White matter microstructure mediates the association between prenatal exposure to phthalates and behavior problems in preschool children. Environmental research 2020; 182: 109093.

46. Vuillermin P, Saffery R, Allen KJ, Carlin JB, Tang ML, Ranganathan S et al. Cohort Profile: The Barwon Infant Study. International Journal of Epidemiology 2015; 44(4): 1148–1160.

47. Adibi JJ, Whyatt RM, Williams PL, Calafat AM, Camann D, Herrick R et al. Characterization of phthalate exposure among pregnant women assessed by repeat air and urine samples. Environ Health Perspect 2008; 116(4): 467–473.

48. Suzuki Y, Niwa M, Yoshinaga J, Watanabe C, Mizumoto Y, Serizawa S et al. Exposure assessment of phthalate esters in Japanese pregnant women by using urinary metabolite analysis. Environmental Health and Preventive Medicine 2009; 14(3): 180–187.

49. Tanner S, Thomson S, Drummond K, O’Hely M, Symeonides C, Mansell T et al. A Pathway-Based Genetic Score for Oxidative Stress: An Indicator of Host Vulnerability to Phthalate-Associated Adverse Neurodevelopment. Antioxidants (Basel) 2022; 11(4): 659.

50. Soininen P, Kangas AJ, Würtz P, Tukiainen T, Tynkkynen T, Laatikainen R et al. High-throughput serum NMR metabonomics for cost-effective holistic studies on systemic metabolism. Analyst 2009; 134(9): 1781–1785.

51. Soininen P, Kangas AJ, Würtz P, Suna T, Ala-Korpela M. Quantitative serum nuclear magnetic resonance metabolomics in cardiovascular epidemiology and genetics. Circ Cardiovasc Genet 2015; 8(1): 192–206.

52. Achenbach TM. DSM-Oriented Guide for the Achenbach System of Empirically Based Assessment (ASEBA). University of Vermont Research Center for Children, Youth, and Families: Burlington, VT, 2013.

53. Goodman R. The Strengths and Difficulties Questionnaire: a research note. Journal of Child Psychology and Psychiatry and Allied Disciplines 1997; 38(5): 581.

54. Pham C, Symeonides C, O’Hely M, Sly PD, Knibbs LD, Thomson S et al. Early life environmental factors associated with autism spectrum disorder symptoms in children at age two years: a birth cohort study. Autism - under review under review.

55. Chericoni N, Balboni G, Costanzo V, Mancini A, Prosperi M, Lasala R et al. A Combined Study on the Use of the Child Behavior Checklist 1½-5 for Identifying Autism Spectrum Disorders at 18 Months. J Autism Dev Disord 2021.

56. Salayev KA, Sanne B. The strengths and difficulties questionnaire (SDQ) in autism spectrum disorders. International Journal on Disability and Human Development 2017; 16(3): 275–280.

57. Jewison T, Su Y, Disfany FM, Liang Y, Knox C, Maciejewski A et al. SMPDB 2.0: big improvements to the Small Molecule Pathway Database. Nucleic acids research 2014; 42(Dx1): D478–D484.

58. Ponsonby A-L. Reflection on modern methods: building causal evidence within high-dimensional molecular epidemiological studies of moderate size. International Journal of Epidemiology 2021; 50(3): 1016–1029.

59. Tingley D, Yamamoto T, Hirose K, Keele L, Imai K. Mediation: R package for causal mediation analysis. 2014.

60. EFSA. Opinion of the Scientific Panel on food additives, flavourings, processing aids and materials in contact with food (AFC) related to Bis (2-ethylhexyl) phthalate (DEHP) for use in food contact materials. EFSA Journal 2005; 3(9): 243.

61. Posnack NG, Swift LM, Kay MW, Lee NH, Sarvazyan N. Phthalate exposure changes the metabolic profile of cardiac muscle cells. Environ Health Perspect 2012; 120(9): 1243–1251.

62. Chiang HC, Kuo YT, Shen CC, Lin YH, Wang SL, Tsou TC. Mono(2-ethylhexyl)phthalate accumulation disturbs energy metabolism of fat cells. Arch Toxicol 2016; 90(3): 589–601.

63. Ellero-Simatos S, Claus SP, Benelli C, Forest C, Letourneur F, Cagnard N et al. Combined transcriptomic-(1)H NMR metabonomic study reveals that monoethylhexyl phthalate stimulates adipogenesis and glyceroneogenesis in human adipocytes. J Proteome Res 2011; 10(12): 5493–5502.

64. Moss EJ, Cook MW, Thomas LV, Gray TJB. The effect of mono-(2-ethylhexyl) phthalate and other phthalate esters on lactate production by Sertoli cells in vitro. Toxicology Letters 1988; 40(1): 77–84.

65. Martinelli MI, Mocchiutti NO, Bernal CA. Dietary di(2-ethylhexyl)phthalate-impaired glucose metabolism in experimental animals. Hum Exp Toxicol 2006; 25(9): 531–538.

66. Kratochvil I, Hofmann T, Rother S, Schlichting R, Moretti R, Scharnweber D et al. MEHP and MEOHP but not DEHP bind productively to the peroxisome proliferator-activated receptor γ. Rapid communications in mass spectrometry: RCM 2019; 33(Suppl 1): 75.

67. Chen Y, McCommis KS, Ferguson D, Hall AM, Harris CA, Finck BN. Inhibition of the Mitochondrial Pyruvate Carrier by Tolylfluanid. Endocrinology 2018; 159(2): 609–621.

68. Rowdhwal SSS, Chen J. Toxic effects of di-2-ethylhexyl phthalate: an overview. BioMed research international 2018; 2018.

69. Liu J, Litt L, Segal MR, Kelly MJ, Pelton JG, Kim M. Metabolomics of oxidative stress in recent studies of endogenous and exogenously administered intermediate metabolites. Int J Mol Sci 2011; 12(10): 6469–6501.

70. Chen YH, Wu YJ, Chen WC, Lee TS, Tsou TC, Chang HC et al. MEHP interferes with mitochondrial functions and homeostasis in skeletal muscle cells. Biosci Rep 2020; 40(4).

71. Li X, Fang EF, Scheibye-Knudsen M, Cui H, Qiu L, Li J et al. Di-(2-ethylhexyl) phthalate inhibits DNA replication leading to hyperPARylation, SIRT1 attenuation, and mitochondrial dysfunction in the testis. Sci Rep 2014; 4: 6434.

72. DeBerardinis RJ, Chandel NS. We need to talk about the Warburg effect. Nat Metab 2020; 2(2): 127–129.

73. Thompson Legault J, Strittmatter L, Tardif J, Sharma R, Tremblay-Vaillancourt V, Aubut C et al. A Metabolic Signature of Mitochondrial Dysfunction Revealed through a Monogenic Form of Leigh Syndrome. Cell Rep 2015; 13(5): 981–989.

74. Begriche K, Massart J, Robin M-A, Borgne-Sanchez A, Fromenty B. Drug-induced toxicity on mitochondria and lipid metabolism: mechanistic diversity and deleterious consequences for the liver. Journal of hepatology 2011; 54(4): 773–794.

75. Xiang AH. Association of Maternal Diabetes With Autism in Offspring. Jama 2017; 317(5): 537–538.

76. Connolly N, Anixt J, Manning P, Ping ILD, Marsolo KA, Bowers K. Maternal metabolic risk factors for autism spectrum disorder-An analysis of electronic medical records and linked birth data. Autism Res 2016; 9(8): 829–837.

77. Hermann R, Lay D, Wahl P, Roth WT, Petrowski K. Effects of psychosocial and physical stress on lactate and anxiety levels. Stress 2019; 22(6): 664–669.

78. Walker CK, Krakowiak P, Baker A, Hansen RL, Ozonoff S, Hertz-Picciotto I. Preeclampsia, placental insufficiency, and autism spectrum disorder or developmental delay. JAMA Pediatr 2015; 169(2): 154–162.

79. Nagalakshmi CS, Santhosh NU, Krishnamurthy N, Chethan C, Shilpashree MK. Role of Altered Venous Blood Lactate and HbA1c in Women with Gestational Diabetes Mellitus. Journal of clinical and diagnostic research : JCDR 2016; 10(12): BC18–BC20.

80. Huo T, Chen X, Lu X, Qu L, Liu Y, Cai S. An effective assessment of valproate sodium-induced hepatotoxicity with UPLC-MS and (1)HNMR-based metabonomics approach. J Chromatogr B Analyt Technol Biomed Life Sci 2014; 969: 109–116.

81. Ottens TH, Nijsten MWN, Hofland J, Dieleman JM, Hoekstra M, van Dijk D et al. Effect of high-dose dexamethasone on perioperative lactate levels and glucose control: a randomized controlled trial. Critical Care 2015; 19(1).

82. Christensen J, Grønborg TK, Sørensen MJ, Schendel D, Parner ET, Pedersen LH et al. Prenatal valproate exposure and risk of autism spectrum disorders and childhood autism. Jama 2013; 309(16): 1696–1703.

83. Namba T, Nardelli J, Gressens P, Huttner WB. Metabolic Regulation of Neocortical Expansion in Development and Evolution. Neuron 2021; 109(3): 408–419.

84. Rash BG, Micali N, Huttner AJ, Morozov YM, Horvath TL, Rakic P. Metabolic regulation and glucose sensitivity of cortical radial glial cells. Proc Natl Acad Sci U S A 2018; 115(40): 10142– 10147.

85. Fernandes DJ, Spring S, Roy AR, Qiu LR, Yee Y, Nieman BJ et al. Exposure to maternal high-fat diet induces extensive changes in the brain of adult offspring. Transl Psychiatry 2021; 11(1): 149.

86. Frye RE, Cakir J, Rose S, Delhey L, Bennuri SC, Tippett M et al. Prenatal air pollution influences neurodevelopment and behavior in autism spectrum disorder by modulating mitochondrial physiology. Mol Psychiatry 2021; 26(5): 1561–1577.

87. Bauer ME, Balistreri M, MacEachern M, Cassidy R, Schoenfeld R, Sankar K et al. Normal Range for Maternal Lactic Acid during Pregnancy and Labor: A Systematic Review and Meta-Analysis of Observational Studies. Am J Perinatol 2019; 36(9): 898–906.

88. Wang Q, Würtz P, Auro K, Mäkinen VP, Kangas AJ, Soininen P et al. Metabolic profiling of pregnancy: cross-sectional and longitudinal evidence. BMC Med 2016; 14(1): 205.

