## Supplement for "Increased maternal non-oxidative energy metabolism mediates association between prenatal DEHP exposure and offspring ASD symptoms: a birth cohort study"

**Article title:**

Table of contents

*Figures*

**Figure 1S. Mediation concept diagram**

**Figure 2S. Results of regression analyses involving prenatal DEHP daily intake, maternal serum metabolites and offspring ASD symptoms**

**Figure 3S. Results of regression analyses for additional prenatal phthalate daily intake measures and maternal serum metabolites**

**Figure 4S. Results of regression analyses involving prenatal DEHP daily intake, cord serum metabolites at birth and offspring ASD symptoms**

**Figure 5S. Results of regression analyses for additional prenatal phthalate daily intake measures and cord serum metabolites**

**Figure 6S. Results of regression analyses involving prenatal DEHP daily intake, child plasma metabolites at 1 year and offspring ASD symptoms**

**Figure 7S. Results of regression analyses for additional prenatal phthalate daily intake measures and child plasma metabolites at 1 year**

**Figure 8S. Results of regression analyses involving prenatal DEHP daily intake, maternal serum metabolites and offspring ASD symptoms stratified by child sex**

**Figure 9S. DEHP exposure may shift metabolism towards inefficient non-oxidative energy metabolism in maternal serum at 28 weeks**

*Tables*

**Table 1S. Names and abbreviations of phthalate compounds and metabolites**

**Table 2S. Additional adjustment factors considered in change-in-estimate analyses**

**Table 3S: Distribution of batch- and SG-corrected phthalate metabolite urine levels (µg/L) and phthalate parent compound daily intake levels (µg/kg bw/day) for 842 mothers**

**Table 4S. Distribution of metabolites in cord serum at birth and child’s plasma at 1 year**

**Table 5S. Results of regression analyses involving prenatal phthalate daily intakes, maternal serum metabolites and offspring ASD symptoms**

*References*

**Figure 1S. Mediation concept diagram**

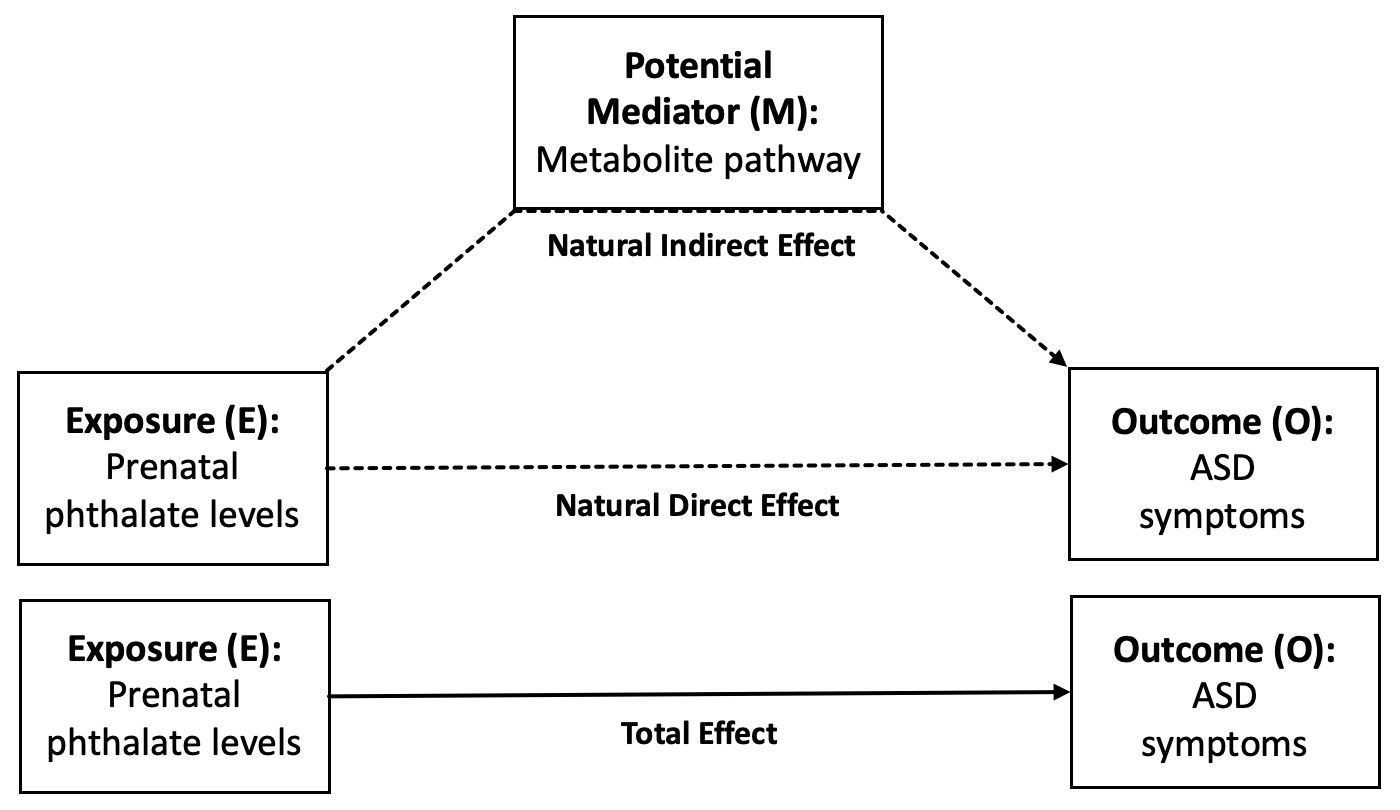

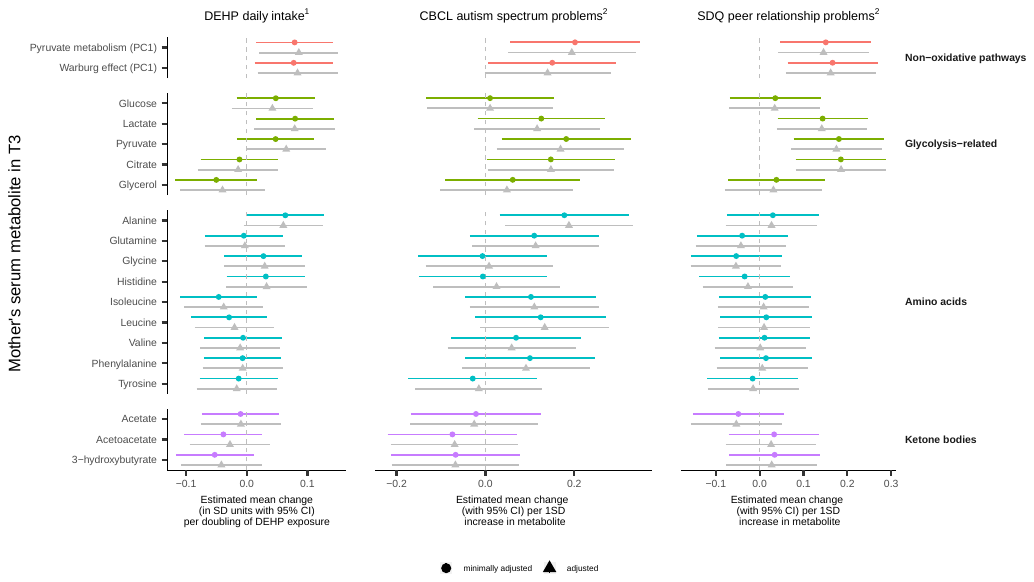

**Figure 2S. Results of regression analyses involving prenatal DEHP daily intake, maternal serum metabolites and offspring ASD symptoms**

DEHP, di(2-ethylhexyl) phthalate; CBCL-ASP, Child Behavior Checklist autism spectrum problems subscale; SDQ-peer, Strengths and Difficulties Questionnaire peer relationship problems subscale; T3, Trimester 3;

^1^ Minimal adjustment set included child’s sex, gestational age at blood collection, gestational age at urine collection; Adjustment set additionally included mother’s age at conception, any maternal smoking in pregnancy, maternal diet in pregnancy;

^2^ Minimal adjustment set included child’s sex, gestational age at blood collection, child’s age at ASD symptom questionnaire; Adjustment set additionally included socioeconomic disadvantage, maternal multiparity.

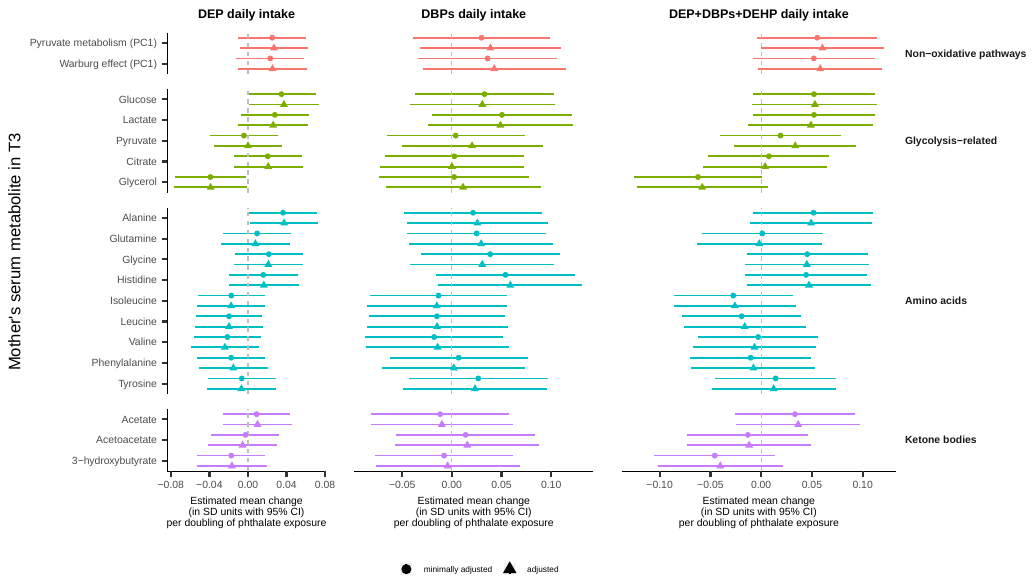

**Figure 3S. Results of regression analyses for additional prenatal phthalate daily intake measures and maternal serum metabolites**

DEHP, di(2-ethylhexyl) phthalate; CBCL-ASP, Child Behavior Checklist autism spectrum problems subscale; SDQ-peer, Strengths and Difficulties Questionnaire peer relationship problems subscale; T3, Trimester 3;

Minimal adjustment set included child’s sex, gestational age at blood collection, gestational age at urine collection; Adjustment set additionally included mother’s age at conception, any maternal smoking in pregnancy, maternal diet in pregnancy.

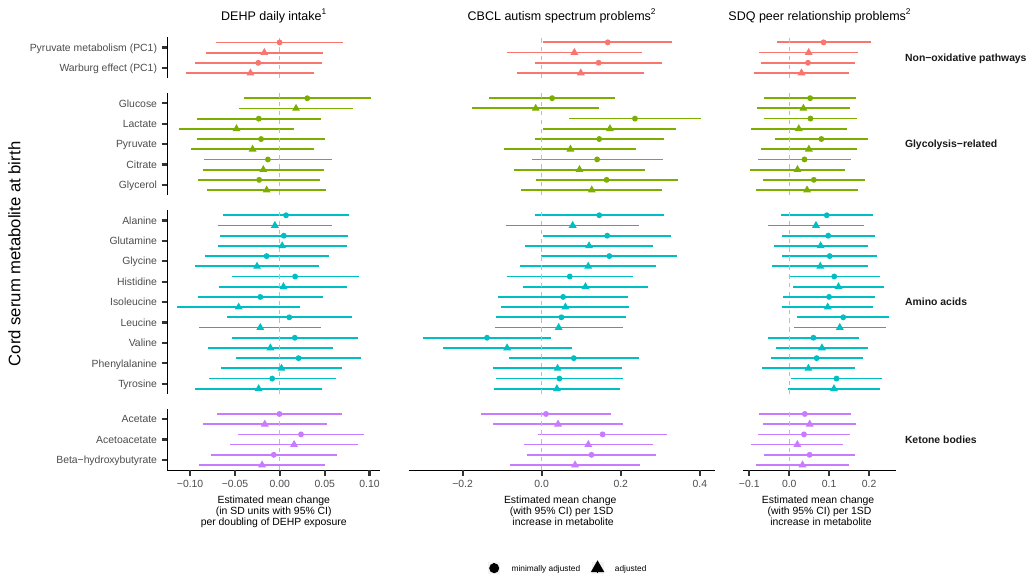

**Figure 4S. Results of regression analyses involving prenatal DEHP daily intake, cord serum metabolites and offspring ASD symptoms**

DEHP, di(2-ethylhexyl) phthalate; CBCL-ASP, Child Behavior Checklist autism spectrum problems subscale; SDQ-peer, Strengths and Difficulties Questionnaire peer relationship problems subscale;

^1^ Minimal adjustment set included child’s sex, gestational age at urine collection, maternal contamination of cord serum; Adjustment set additionally included mother’s age at conception, any maternal smoking in pregnancy, Apgar score at 1 minute, mode of birth;

^2^ Minimal adjustment set included child’s sex, maternal contamination of cord serum, child’s age at ASD symptom questionnaire; Adjustment set additionally included socioeconomic disadvantage, maternal multiparity.

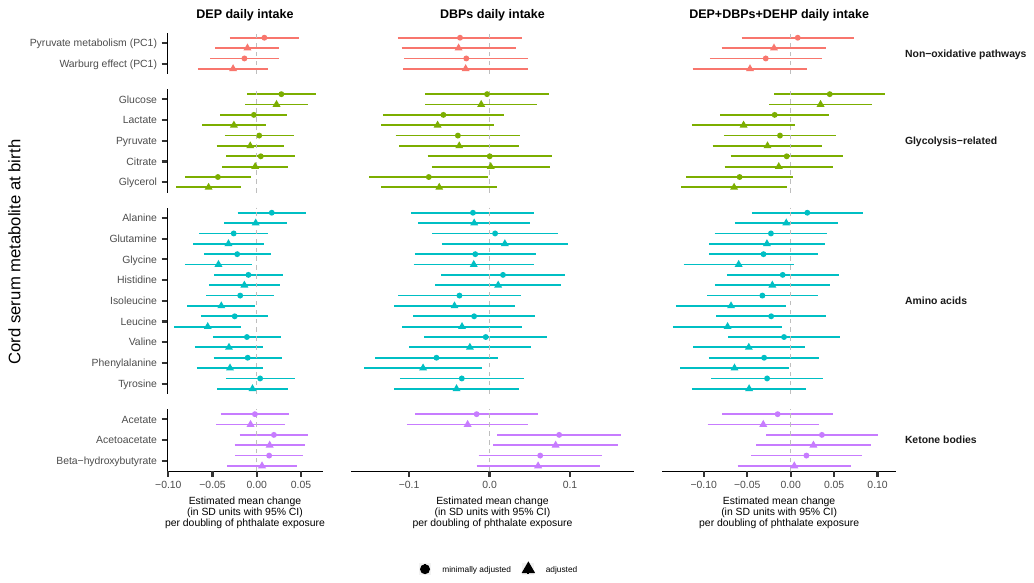

**Figure 5S. Results of regression analyses for additional prenatal phthalate daily intake measures and cord serum metabolites**

DEHP, di(2-ethylhexyl) phthalate; CBCL-ASP, Child Behavior Checklist autism spectrum problems subscale; SDQ-peer, Strengths and Difficulties Questionnaire peer relationship problems subscale;

Minimal adjustment set included child’s sex, gestational age at urine collection, maternal contamination of cord serum; Adjustment set additionally included mother’s age at conception, any maternal smoking in pregnancy, Apgar score at 1 minute, mode of birth.

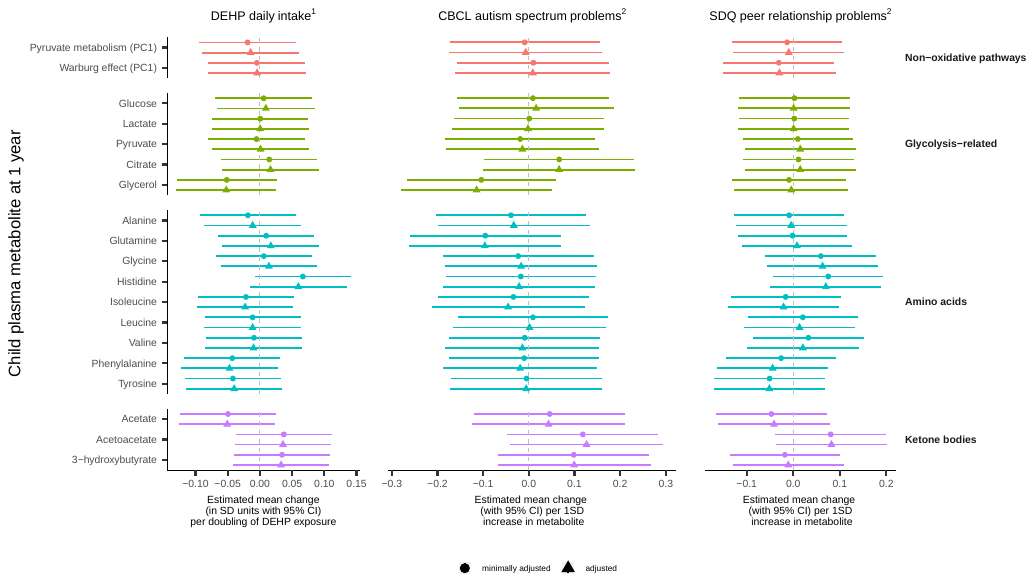
 **Figure 6S. Results of regression analyses involving prenatal DEHP daily intake, child plasma metabolites at 1 year and offspring ASD symptoms**

DEHP, di(2-ethylhexyl) phthalate; CBCL-ASP, Child Behavior Checklist autism spectrum problems subscale; SDQ-peer, Strengths and Difficulties Questionnaire peer relationship problems subscale;

^1^ Minimal adjustment set included child’s sex, gestational age at urine collection, child’s age at blood collection; Adjustment set additionally included mother’s age at conception, any maternal smoking in pregnancy, child’s weight at 1 year of age, season as measured by ultraviolet radiation at 9 months;

^2^ Minimal adjustment set included child’s sex, child’s age at blood collection, child’s age at ASD symptom questionnaire; Adjustment set additionally included socioeconomic disadvantage, maternal multiparity.

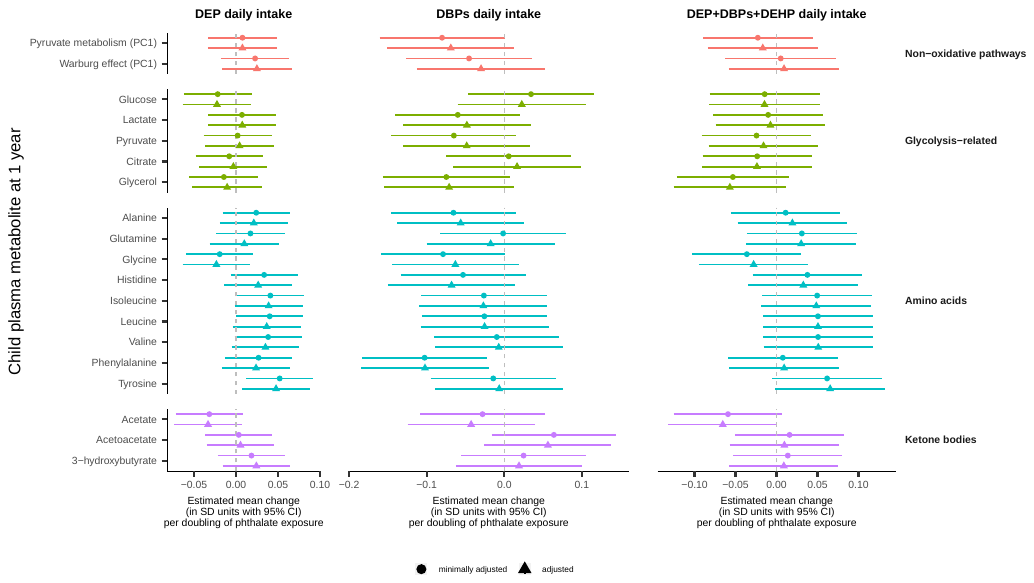

**Figure 7S. Results of regression analyses for additional prenatal phthalate daily intake measures and child plasma metabolites at 1 year**

DEHP, di(2-ethylhexyl) phthalate; CBCL-ASP, Child Behavior Checklist autism spectrum problems subscale; SDQ-peer, Strengths and Difficulties Questionnaire peer relationship problems subscale;

Minimal adjustment set included child’s sex, gestational age at urine collection, child’s age at blood collection; Adjustment set additionally included mother’s age at conception, any maternal smoking in pregnancy, child’s weight at 1 year of age, season as measured by ultraviolet radiation at 9 months.

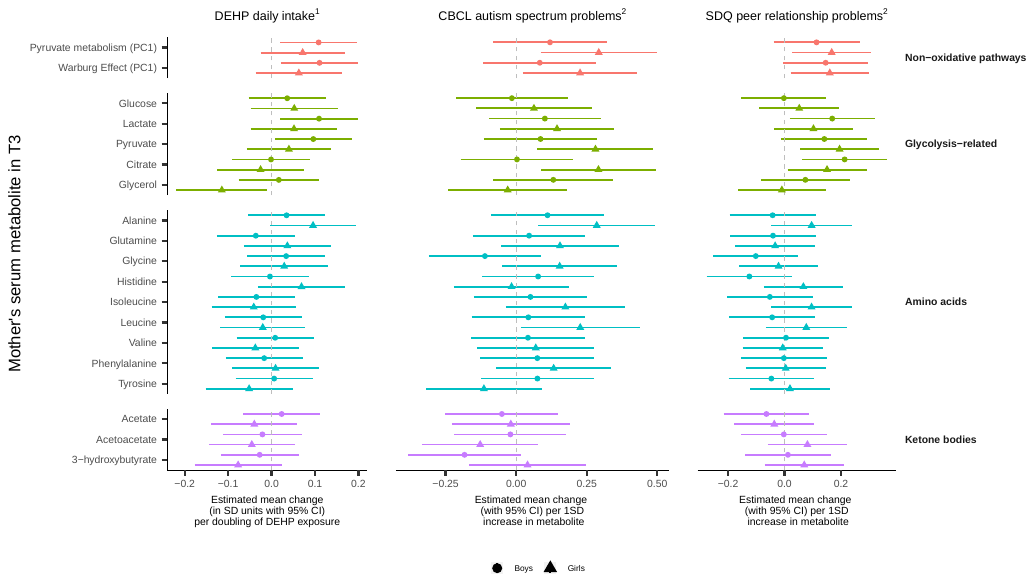
 **Figure 8S. Results of regression analyses involving prenatal DEHP daily intake, maternal serum metabolites and offspring ASD symptoms stratified by child sex**

DEHP, di(2-ethylhexyl) phthalate; CBCL-ASP, Child Behavior Checklist autism spectrum problems subscale; SDQ-peer, Strengths and Difficulties Questionnaire peer relationship problems subscale; T3, Trimester 3;

^1^ Model adjusted for child’s sex, gestational age at blood collection, gestational age at urine collection, mother’s age at conception, any maternal smoking in pregnancy, maternal diet in pregnancy;

^2^ Model adjusted for child’s sex, gestational age at blood collection, child’s age at ASD symptom questionnaire, socioeconomic disadvantage, maternal multiparity.

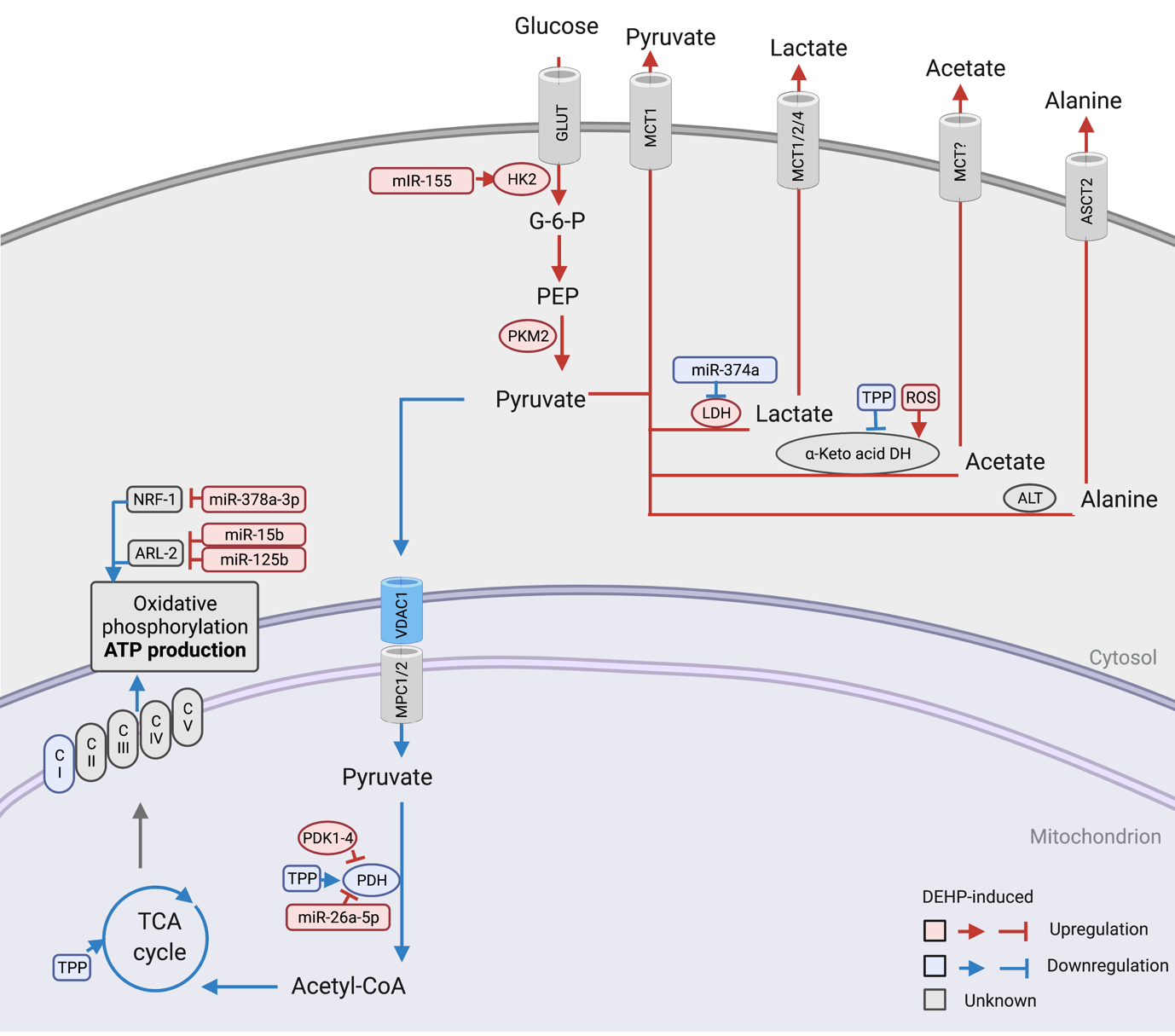

**Created with BioRender.com**

**Figure 9S.** **DEHP exposure may shift metabolism towards inefficient non-oxidative energy metabolism in maternal serum at 28 weeks.** Pyruvate is the end-product of glycolysis and can undergo oxidative and non-oxidative metabolism. Pyruvate oxidation typically occurs when pyruvate is transported into mitochondria and into the tricarboxylic acid (TCA) cycle to drive efficient ATP production via oxidative phosphorylation. This relies on mitochondrial transporters (Voltage-Dependent Anion Channel 1 (VDAC1) and mitochondrial pyruvate carrier (MPC1/2)) to uptake cytosolic pyruvate into the mitochondria and maintain mitochondrial pyruvate flux ^1^. Once in the mitochondria, pyruvate is oxidized into acetyl CoA via decarboxylation with the enzyme pyruvate dehydrogenase (PDH). Under certain conditions, like hyperactive cellular metabolism or mitochondria dysfunction, excess carbon from glycolysis could lead to an elevation of pyruvate and non-oxidative by-products of pyruvate metabolism (i.e., lactate, acetate and alanine). The conversion of pyruvate to lactate by lactate dehydrogenase (LDH) occurs under these conditions (aerobic glycolysis, i.e., the Warburg Effect).^2^ The conversion of pyruvate to alanine by alanine aminotransferase (ALT) also can occur.^3^ The conversion of pyruvate to acetate by pyruvate oxidase (POXB) also can occur under conditions of nutrient deficiency or oxidative stress.^4, 5^ Excess cytosolic pyruvate can be exported via monocarboxylate transporter 1 (MCT1). Phthalate exposure could *directly* result in a metabolic shift that elevates non-oxidative energy metabolism and the Warburg effect pathway. This could occur through multiple known or potentional actions of phthalates (e.g., disrupted mitochondrial pyruvate flux (VDAC1 and MPC), mitochondrial PDH activity, increased dimeric PKM2, elevated basal glucose uptake or increased activity of glycolytic enzymes^2, 6^). The metabolic shift may also be a result of *indirect* effects of phthalate exposure, like mitochondrial dysfunction or oxidative stress. A metabolic shift may also lead to increased oxidative stress due to the generation of reactive oxidative stress (ROS) products^7-10^ and their reduced elimination.^11^ This could positively feed-back to exacerbate any underlying mitochondrial dysfunction.^12^

**Table 1S. Names and abbreviations of phthalate compounds and metabolites**

| **Parent compound** | | **Urinary metabolite** | |
| --- | --- | --- | --- |
| Abbreviation | Full name | Abbreviation | Full name |
| DEP | Diethyl phthalate | MEP | Monoethyl phthalate |
| DiBP | Diisobutyl phthalate | MiBP | Monoisobutyl phthalate |
| DnBP | Di-n-butyl phthalate | MnBP | Mono-n-butyl phthalate |
| DEHP | Di-(2-ethylhexyl) phthalate | MEHP | Mono-(2-ethylhexyl) phthalate |
|  |  | MEHHP | Mono-(2-ethyl-5-hydroxyhexyl) phthalate |
|  |  | MECPP | Mono-(5-carboxy-2-ethylpentyl) phthalate |
|  |  | MEOHP | Mono-(2-ethyl-5-oxohexyl) phthalate |

**Table 2S. Additional adjustment factors considered in change-in-estimate analyses**

The following factors were individually added to the multivariable regression models at each metabolomic time point (unless specified in brackets), and their inclusion did not materially change the results.

| Regression of metabolite on prenatal phthalate daily intake | | Regression of ASD symptoms on metabolite | |
| --- | --- | --- | --- |
| ALL METABOLITE TIME POINTS | SELECT METABOLITE TIME POINTS | ALL METABOLITE TIME POINTS | SELECT METABOLITE TIME POINTS |
| - Maternal multiparity - Maternal education - Maternal alcohol use in pregnancy - Maternal perceived stress in pregnancy | - Season as measured by ultraviolet radiation in trimester 1 *(prenatal, birth)* | - Maternal age at conception - Maternal alcohol use in pregnancy - Maternal fish oil supplementation in pregnancy - Air pollution | - Parent country of birth *(postnatal)* - Maternal perceived stress at 6 months *(postnatal)* - Childcare use prior to 2 years *(postnatal)* |

**Table 3S:** **Distribution of batch- and SG-corrected phthalate metabolite urine levels (µg/L) and phthalate parent compound daily intake levels (µg/kg bw/day) for 842 mothers**

| Parent | Metabolite | LOD | %>LOD | GM (95% CI) | 25^th^ percentile | Median | 75^th^ percentile | Range | GM daily intake levels (95% CI)^a^ |
| --- | --- | --- | --- | --- | --- | --- | --- | --- | --- |
| DEP | MEP | 0.34 | 99 | 53.5 (48.9, 58.5) | 22.8 | 45.5 | 108.5 | <LOD-7558 | 1.6 (1.4, 1.7) |
| DBPs | MiBP | 3.9 | 98 | 17.8 (16.9, 18.7) | 11.1 | 17.4 | 27.4 | <LOD-451 | 1.9 (1.8, 2.0)^b^ |
|  | MnBP | 4.5 | 99 | 21.7 (20.7, 22.7) | 13.6 | 21.3 | 33.8 | <LOD-248 |  |
| DEHP | MEHP^c^ | 4.1 | 33 | 4.0 (3.8, 4.1) | <LOD | <LOD | 4.9 | <LOD-1032 | 1.6 (1.5, 1.7)^d^ |
|  | MEHHP | 0.13 | 100 | 9.8 (9.3, 10.3) | 6.3 | 9.7 | 14.9 | 0.3-2709 |  |
|  | MECPP | 0.03 | 100 | 12.7 (12.1, 13.3) | 8.5 | 12.1 | 18.0 | 1.1-1726 |  |
|  | MEOHP | 0.10 | 100 | 8.2 (7.8, 8.6) | 5.4 | 8.1 | 12.3 | 0.3-1365 |  |

*NB.* SG, specific gravity; LOD, limit of detection (µg/L); GM, geometric mean; CI, confidence interval; DEP, Diethyl phthalate; DBPs, sum of DiBP (Diisobutyl phthalate) and DnBP (Di-n-butyl phthalate); DEHP, Di-(2-ethylhexyl) phthalate; MEP, Monoethyl phthalate; MiBP, Monoisobutyl phthalate; MnBP, Mono-n-butyl phthalate; MEHP, Mono(2-ethylhexyl) phthalate; MEHHP, Mono(2-ethyl-5-hydroxyhexyl) phthalate; MECPP, Mono(5-carboxy-2-ethylpentyl) phthalate; MEOHP, Mono-(2-ethyl-5-oxohexyl) phthalate;

^a^Includes standardization for time of day of urine collection;

^b^GM daily intake for DBPs calculated from the sum of MiBP and MnBP;

^c^Not batch corrected due to contamination;

^d^GM daily intake for DEHP calculated from the sum of MEHHP, MECPP and MEOHP.

**Table 4S. Distribution of metabolites in cord serum at birth and child’s plasma at 1 year**

|  | **Full sample** | | |
| --- | --- | --- | --- |
|  | N | | Mean (SD) |
| **Cord serum glycolysis-related metabolites** | | | |
| Glucose ($\mu$mol/L) | 907 | 3037.9 (1071.1) | |
| Lactate ($\mu$mol/L) | 909 | 4980.9 (2148.7) | |
| Pyruvate ($\mu$mol/L) | 881 | 189.6 (93.1) | |
| Citrate ($\mu$mol/L) | 884 | 117.8 (26.5) | |
| Glycerol ($\mu$mol/L) | 864 | 90.9 (74.6) | |
| **Cord serum amino acids** | | | |
| Alanine ($\mu$mol/L) | 909 | 445.0 (89.3) | |
| Glutamine ($\mu$mol/L) | 882 | 462.1 (84.9) | |
| Glycine ($\mu$mol/L) | 899 | 293.4 (69.8) | |
| Histidine ($\mu$mol/L) | 907 | 83.7 (15.1) | |
| Isoleucine ($\mu$mol/L) | 909 | 48.3 (11.2) | |
| Leucine ($\mu$mol/L) | 909 | 67.5 (16.7) | |
| Valine ($\mu$mol/L) | 906 | 165.5 (27.5) | |
| Phenylalanine ($\mu$mol/L) | 909 | 89.8 (14.4) | |
| Tyrosine ($\mu$mol/L) | 906 | 57.5 (12.3) | |
| **Cord serum ketone bodies** | | | |
| Acetate ($\mu$mol/L) | 909 | 46.8 (8.9) | |
| Acetoacetate ($\mu$mol/L) | 909 | 52.5 (40.1) | |
| 3-hydroxybutyrate ($\mu$mol/L) | 907 | 275.8 (160.1) | |
| **1 year plasma glycolysis-related metabolites** | | | |
| Glucose ($\mu$mol/L) | 729 | 2841.8 (1044.4) | |
| Lactate ($\mu$mol/L) | 742 | 3051.8 (2148.2) | |
| Pyruvate ($\mu$mol/L) | 739 | 196.5 (137.9) | |
| Citrate ($\mu$mol/L) | 742 | 124.8 (18.5) | |
| Glycerol ($\mu$mol/L) | 702 | 110.1 (38.1) | |
| **1 year plasma amino acids** | | | |
| Alanine ($\mu$mol/L) | 742 | 370.7 (79.2) | |
| Glutamine ($\mu$mol/L) | 740 | 435.9 (62.7) | |
| Glycine ($\mu$mol/L) | 742 | 228.8 (41.1) | |
| Histidine ($\mu$mol/L) | 741 | 66.5 (14.1) | |
| Isoleucine ($\mu$mol/L) | 742 | 74.1 (24.8) | |
| Leucine ($\mu$mol/L) | 742 | 91.3 (31.1) | |
| Valine ($\mu$mol/L) | 742 | 208.9 (68.4) | |
| Phenylalanine ($\mu$mol/L) | 742 | 66.5 (14.7) | |
| Tyrosine ($\mu$mol/L) | 740 | 71.7 (25.4) | |
| **1 year plasma ketone bodies** | | | |
| Acetate ($\mu$mol/L) | 742 | 39.9 (15.0) | |
| Acetoacetate ($\mu$mol/L) | 742 | 71.2 (62.4) | |
| 3-hydroxybutyrate ($\mu$mol/L) | 742 | 187.7 (148.9) | |

*NB.* SD, standard deviation.

**Table 5S. Results of regression analyses involving prenatal DEHP daily intake, maternal serum metabolites and offspring ASD symptoms**

|  | **DEHP daily intake^a^** | | **CBCL autism spectrum problems^b^** | | **SDQ peer relationship problems^b^** | |
| --- | --- | --- | --- | --- | --- | --- |
| **Metabolite (**$\boldsymbol{\mu}$**mol/L)** | $\beta$ (95% CI)^c^ | *p*-value | $\beta$ (95% CI)^d^ | *p*-value | $\beta$ (95% CI)^d^ | *p*-value |
| **Non-oxidative pathways** | | | | | | |
| Pyruvate metabolism (PC1) | 113.32 (27.47, 199.18) | 0.01 | 0.14 (0.04, 0.24) | 0.008 | 0.11 (0.03, 0.18) | 0.006 |
| Warburg Effect (PC1) | 108.25 (22.93, 193.56) | 0.01 | 0.10 (0.00, 0.21) | 0.05 | 0.12 (0.04, 0.20) | 0.002 |
| **Glycolysis-related** | | | | | | |
| Glucose | 54.15 (-31.30, 139.61) | 0.21 | 0.01 (-0.10, 0.12) | 0.88 | 0.03 (-0.05, 0.11) | 0.51 |
| Lactate | 38.68 (6.09, 71.28) | 0.02 | 0.23 (-0.05, 0.51) | 0.11 | 0.28 (0.08, 0.48) | 0.007 |
| Pyruvate | 2.37 (0.00, 4.74) | 0.05 | 4.42 (0.67, 8.17) | 0.02 | 4.74 (1.94, 7.53) | 0.001 |
| Citrate | -0.27 (-1.53, 0.99) | 0.67 | 7.83 (0.28, 15.39) | 0.04 | 10.04 (4.48, 15.60) | 0.0004 |
| Glycerol | -1.24 (-3.40, 0.91) | 0.26 | 1.56 (-3.27, 6.40) | 0.53 | 1.03 (-2.58, 4.63) | 0.58 |
| **Amino acids** | | | | | | |
| Alanine | 2.77 (-0.23, 5.76) | 0.07 | 4.06 (0.95, 7.18) | 0.01 | 0.58 (-1.67, 2.84) | 0.61 |
| Glutamine | -0.17 (-3.85, 3.51) | 0.93 | 1.99 (-0.52, 4.49) | 0.12 | -0.77 (-2.62, 1.09) | 0.42 |
| Glycine | 1.02 (-1.27, 3.31) | 0.38 | 0.24 (-3.88, 4.36) | 0.91 | -1.55 (-4.52, 1.42) | 0.31 |
| Histidine | 0.45 (-0.47, 1.37) | 0.34 | 1.85 (-8.66, 12.37) | 0.73 | -1.97 (-9.65, 5.70) | 0.61 |
| Isoleucine | -0.58 (-1.56, 0.41) | 0.25 | 7.34 (-2.33, 17.00) | 0.14 | 0.60 (-6.30, 7.50) | 0.87 |
| Leucine | -0.32 (-1.36, 0.72) | 0.55 | 8.55 (-0.74, 17.84) | 0.07 | 0.65 (-6.11, 7.42) | 0.85 |
| Valine | -0.32 (-2.24, 1.61) | 0.75 | 1.96 (-2.82, 6.75) | 0.42 | 0.05 (-3.52, 3.62) | 0.98 |
| Phenylalanine | -0.10 (-1.07, 0.87) | 0.84 | 6.19 (-3.51, 15.89) | 0.21 | 0.41 (-6.63, 7.44) | 0.91 |
| Tyrosine | -0.18 (-0.91, 0.55) | 0.63 | -1.35 (-14.47, 11.76) | 0.84 | -1.36 (-10.84, 8.12) | 0.78 |
| **Ketone bodies** | | | | | | |
| Acetate | -0.09 (-0.67, 0.50) | 0.78 | -2.71 (-18.23, 12.81) | 0.73 | -5.78 (-17.04, 5.48) | 0.31 |
| Acetoacetate | -0.52 (-1.78, 0.73) | 0.41 | -6.21 (-19.11, 6.68) | 0.34 | 1.34 (-3.95, 6.63) | 0.62 |
| 3-hydroxybutyrate | -2.71 (-7.03, 1.61) | 0.22 | -1.74 (-5.42, 1.94) | 0.35 | 0.41 (-1.14, 1.96) | 0.60 |

*NB.* DEHP, di(2-ethylhexyl) phthalate; CBCL, Child Behavior Checklist; SDQ, Strengths and Difficulties Questionnaire; PC1, principal component 1;

^a^ Model adjusted for child’s sex, gestational age at blood collection, gestational age at urine collection, mother’s age at conception, any maternal smoking in pregnancy, maternal diet in pregnancy;

^b^ Model adjusted for child’s sex, gestational age at blood collection, child’s age at ASD symptom questionnaire, socioeconomic disadvantage, maternal multiparity;

^c^ Estimated increase in metabolite (*μ*mol/L) per doubling of prenatal DEHP daily intake;

^d^ Estimated increase in ASD symptom score per 1,000 $\mu$mol/L increase in metabolite measure.

**References**

1.	Rossi A, Rigotto G, Valente G, Giorgio V, Basso E, Filadi R *et al.* Defective Mitochondrial Pyruvate Flux Affects Cell Bioenergetics in Alzheimer's Disease-Related Models. *Cell Rep* 2020; **30**(7)**:** 2332-2348.e2310.

2.	Liberti MV, Locasale JW. The Warburg Effect: How Does it Benefit Cancer Cells? *Trends Biochem Sci* 2016; **41**(3)**:** 211-218.

3.	Sousa CM, Biancur DE, Wang X, Halbrook CJ, Sherman MH, Zhang L *et al.* Pancreatic stellate cells support tumour metabolism through autophagic alanine secretion. *Nature* 2016; **536**(7617)**:** 479-483.

4.	Bose S, Ramesh V, Locasale JW. Acetate Metabolism in Physiology, Cancer, and Beyond. *Trends Cell Biol* 2019; **29**(9)**:** 695-703.

5.	Liu X, Cooper DE, Cluntun AA, Warmoes MO, Zhao S, Reid MA *et al.* Acetate Production from Glucose and Coupling to Mitochondrial Metabolism in Mammals. *Cell* 2018; **175**(2)**:** 502-513.e513.

6.	Tanner LB, Goglia AG, Wei MH, Sehgal T, Parsons LR, Park JO *et al.* Four Key Steps Control Glycolytic Flux in Mammalian Cells. *Cell Syst* 2018; **7**(1)**:** 49-62.e48.

7.	Yang C, Ko B, Hensley CT, Jiang L, Wasti AT, Kim J *et al.* Glutamine oxidation maintains the TCA cycle and cell survival during impaired mitochondrial pyruvate transport. *Mol Cell* 2014; **56**(3)**:** 414-424.

8.	St-Pierre J, Buckingham JA, Roebuck SJ, Brand MD. Topology of superoxide production from different sites in the mitochondrial electron transport chain. *J Biol Chem* 2002; **277**(47)**:** 44784-44790.

9.	Kakimoto PA, Tamaki FK, Cardoso AR, Marana SR, Kowaltowski AJ. H2O2 release from the very long chain acyl-CoA dehydrogenase. *Redox Biol* 2015; **4:** 375-380.

10.	Schönfeld P, Wojtczak L. Fatty acids as modulators of the cellular production of reactive oxygen species. *Free Radic Biol Med* 2008; **45**(3)**:** 231-241.

11.	Liu J, Litt L, Segal MR, Kelly MJ, Pelton JG, Kim M. Metabolomics of oxidative stress in recent studies of endogenous and exogenously administered intermediate metabolites. *Int J Mol Sci* 2011; **12**(10)**:** 6469-6501.

12.	Marí M, Morales A, Colell A, García-Ruiz C, Fernández-Checa JC. Mitochondrial glutathione, a key survival antioxidant. *Antioxid Redox Signal* 2009; **11**(11)**:** 2685-2700.
